# Splitting Schizophrenia: Divergent Cognitive and Educational Outcomes Revealed by Genomic Structural Equation Modelling

**DOI:** 10.1101/2024.10.23.24315121

**Authors:** Cameron James Watson, Johan Zvrskovec, Giuseppe Pierpaolo Merola, Lachlan Gilchrist, Senta M Haussler, Miryam Schattner, Chris Wai Hang Lo, Gerome Breen, Robin M Murray, Cathryn M Lewis, Evangelos Vassos

## Abstract

The genetic relationship between schizophrenia, IQ, and educational attainment (EA) is complex. Schizophrenia polygenic scores (PGS) are linked to lower IQ, whilst higher IQ-PGS correlates with reduced schizophrenia risk. Paradoxically, genetic predisposition to higher EA has been associated with increased schizophrenia risk, a relationship potentially confounded by genetic overlap between schizophrenia and bipolar disorder. Through Genomic Structural Equation Modelling we dissected the genetic contribution to schizophrenia, identifying 63 SNPs uniquely associated with schizophrenia (SZspecific) and 78 shared with bipolar disorder (PSYshared). Both schizophrenia (rg = -0.22) and SZspecific (rg = -0.24) were genetically negatively correlated with IQ, the correlations between bipolar disorder and PSYshared with IQ were less pronounced (both rg = -0.07). Schizophrenia exhibited minimal correlation with EA (rg = 0.01), yet the latent variables demonstrated divergent relationships; PSYshared was positively correlated (rg = 0.11), whereas SZspecific was negatively correlated (rg = -0.06). PGS analyses in the UK Biobank (n=381,688), corroborated these divergent relationships, SZspecific-PGS was negatively associated with EA (β = -0.13, p < 2e-16), whereas the PSYshared-PGS was positively associated (β = 0.14, p < 2e-16). Mendelian Randomisation provided additional support, but also confirmed the presence of genetic pleiotropy. Our findings underscore the utility of genetic methods in dissecting neuropsychiatric disorders, supporting the existence of two possible pathways to schizophrenia: one shared with bipolar disorder and another with stronger neurodevelopmental underpinnings.

## Introduction

Schizophrenia is a disabling condition, with patients losing the equivalent of 73% of healthy life each year (1). With employment levels as low as 10% and a peak incidence in the third decade of life, schizophrenia imposes a major burden on patients, families and society (2). The disorder is complex and characterised by clusters of specific symptoms, including positive symptoms such as hallucinations and delusions, negative symptoms such as affective flattening and avolition, as well as cognitive deficits. Positive symptoms are thought to be driven by striatal dopaminergic excess, and though effectively treated in many cases by antipsychotics, the latter can result in unpleasant side effects (3). In contrast, negative and cognitive symptoms remain largely unaddressed by current pharmacological treatments and their mechanisms are incompletely understood (4, 5).

Cognitive impairment is common in schizophrenia and can appear years before the onset of psychosis (6). Individuals with the disorder score around 1-2 standard deviations below that of healthy controls across a range of neuropsychological tasks (4, 7); major deficits are seen in processing speed, working memory, and new learning (8). Cognitive impairment is also one of the strongest predictors of poor functional outcomes, reducing employment prospects, and the ability to live independently (9). A key focus of research in the past decade has been to delineate whether cognitive impairment is a risk factor for the development of schizophrenia, a premorbid manifestation of the disorder, or both.

Like schizophrenia, with its heritability between 65-80% (10, 11), the twin heritability of cognition is substantial, with estimates ranging between 40-80% (12). Schizophrenia and cognition also share considerable genetic links (13); structural equation modelling has demonstrated that a third of schizophrenia liability indexed by polygenic scores (PGS) is mediated through influences on cognition (14). Mendelian randomization (MR) studies have supported a bidirectional causal relationship between intelligence and schizophrenia, demonstrating that higher intelligence may be protective against diagnosis, whilst schizophrenia is associated with lower levels of IQ (15). Schizophrenia PGS (SZ-PGS) has also been associated with poorer performance in several domains of intelligence (16), whereas the PGS for childhood intelligence was significantly lower in those with schizophrenia than controls (17). Additionally, rare genetic variants that have been robustly associated with schizophrenia are enriched in individuals with autism and intellectual disability (18), supporting the current conceptualisation of these disorders falling on a neurodevelopmental continuum (19).

Despite the consistent negative association between schizophrenia and cognition across a range of study designs, counterintuitive findings have emerged for educational attainment (EA). Large genome-wide association studies (GWAS) have demonstrated significant positive genetic correlations between schizophrenia and EA (16, 20). Liability to higher EA has also been associated with an increased risk of schizophrenia (16), whilst SZ-PGS is associated with increased years in education (21). This paradox is intriguing given that phenotypically schizophrenia and EA appear to be negatively correlated; lower EA in patients is seen in comparison to both controls (22) and healthy siblings (23). Education is also highly correlated with intelligence, both from a genetic (rg = 0.7) (24) and phenotypic (r = 0.8) (25) standpoint. Moreover these traits are highly influential on each other, as intelligence increases EA (20), and vice versa, each year in education increases IQ between 1-5 points (26). In the context of such counterintuitive findings, diverse methods have been employed to untangle the relationship between schizophrenia and EA. Multivariate MR studies have revealed important roles for income and other socioeconomic factors in mediating the relationship (27). PGS studies have yielded mixed results, with some suggesting that improved EA might be driven by non-cognitive phenotypes associated with the genetic liability to schizophrenia. One such example is creativity, which has been associated with PGS for both schizophrenia and bipolar disorder (21). Bipolar disorder, a mood disorder characterised by episodes of mania and depression, has historically been considered distinct from schizophrenia. However, these disorders share significant genotypic and phenotypic overlap (28). It has recently been suggested that the relationship between schizophrenia and EA is mediated by shared genetic risk with bipolar disorder (29), underscoring the importance of dissecting genetic interactions between psychotic disorders.

Using Genomic Structural Equation Modelling (Genomic SEM) (30), we aim to build on these findings and estimate the effect of single nucleotide polymorphisms (SNPs) on schizophrenia independent of their effect on bipolar disorder. This resulting “GWAS-by-subtraction” has previously been employed to segregate the contribution of non-cognitive skills from cognitive skills on educational attainment (31). Here, this technique will allow the identification of SNPs uniquely associated with schizophrenia (SZ_specific_) as opposed to those which are linked to a broader liability to psychosis (PSY_shared_), indexing risk shared between schizophrenia and bipolar disorder. We hypothesise that there will be divergent effects on educational attainment and cognition between SZ_specific_ and PSY_shared_ variables. If this hypothesis were to hold, in-keeping with the neurodevelopmental continuum of psychosis, genetic risk unique to schizophrenia will have a more deleterious impact on EA and cognition, than the risk shared with bipolar disorder.

## Methods

### GWAS-by-subtraction

Following pre-processing of GWAS summary data for schizophrenia (32) and bipolar disorder (33), we conducted a GWAS-by-subtraction. Using methods proposed by Demange et al 2021 (31) (see <u>Supplemental Note</u>), we leveraged existing Genomic SEM principles to estimate the unique effect of each single nucleotide polymorphism (SNP) on schizophrenia, accounting for its effect on bipolar disorder. Our model (Figure 1) resulted in one path representing the genetic effects shared between both schizophrenia and bipolar disorder (PSY_shared_), while the other path measured the SNP effects unique to schizophrenia (SZ_specific_), independent of shared effects. Using PLINK v1.9 (34) we identified independently associated SNPs for SZ_specific_ and PSY_shared_. Based on the genome wide significance threshold of p < 5 × 10^−8^, each significant SNP was also compared to previously published GWAS, including schizophrenia and bipolar disorder, and considered novel if it had not previously been reported in the GWAS Catalog (35).

**Figure 1:**
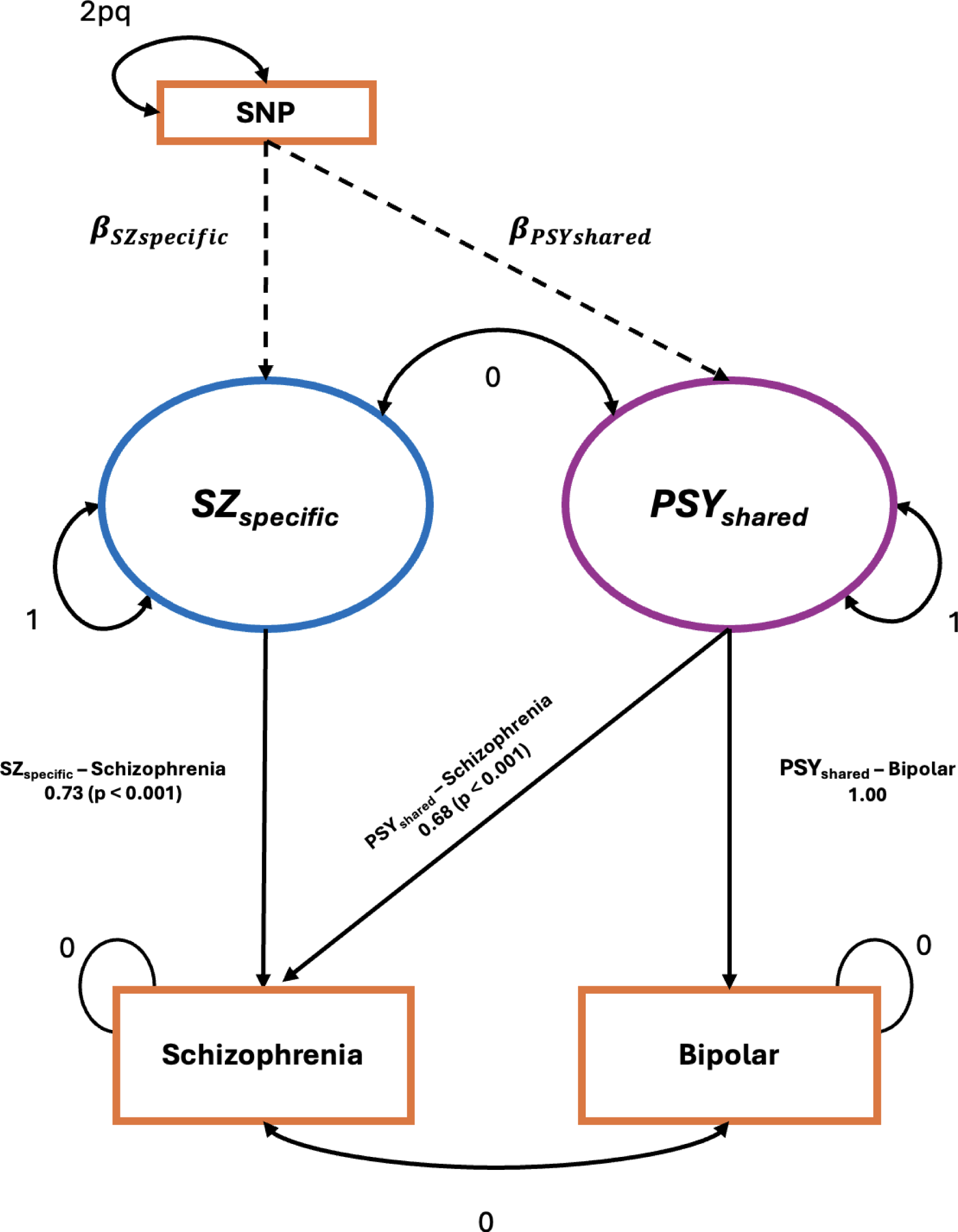
GWAS-by Subtraction model in Genomic SEM with path diagram. Standardised effect sizes and p-values are shown. SNP (single nucleotide polymorphism). SZ_specific_ refers to genetic influences on schizophrenia that are unique to schizophrenia, whereas PSY_shared_ captures genetic influences on schizophrenia that are shared between bipolar disorder and schizophrenia.

### Genetic correlations

The LDSC function in the Genomic SEM package for R (30) was used to calculate the genetic correlation between schizophrenia, bipolar disorder, SZ_specific_, PSY_shared_, educational attainment and IQ, based on GWAS summary statistics. P-values were calculated for each pair, testing the hypothesis that the estimates are different from zero. A Benjamini-Hochberg False Discovery Rate (FDR) was applied to these p-values (q-value = 0.05) to mitigate for multiple testing bias. For full parameters and GWAS summary statistic preparation, see <u>Supplemental Note</u>.

### Polygenic Score Analysis in the UK Biobank

To assess the impact of genetic risk related to psychotic disorders on educational and cognitive traits, we leveraged data from the UK Biobank (Project 82087). The UK Biobank is a large-scale biomedical database and research resource containing genetic and phenotypic data on half a million UK participants (36). At baseline visits (between 2006 - 2010), touchscreen questionnaires were used to gather information on sociodemographic, cognitive and health variables. In 2016, 157,366 participants completed an online Mental Health Questionnaire (MHQ) (37).

#### A) Polygenic score creation

Polygenic scores were created based on summary data from schizophrenia (32) and bipolar disorder GWAS (33) and the GWAS of our latent variables, SZ_specific_ and PSY_shared_. PGS were calculated using MegaPRS (38), leveraging the GenoPred pipeline (39). After ensuring the quality of genetic data and addressing potential relatedness between participants (see <u>Supplemental Note</u>) the resulting cohort consisted of 381,688 individuals.

#### B) Educational attainment (EA)

Educational attainment (EA) (<u>ID: 6138</u>) was assessed via touchscreen questionnaire, with participants reporting their highest level of qualification across four possible study visits. Replicating methods implemented in recent large studies (40), we derived an Education Years estimate based on the average number of years in education associated with specific qualifications (see <u>Supplemental Note</u>). Linear regression models were used to examine the relationship between Education Years and polygenic scores (SZ-PGS, BD-PGS, SZ_specific_-PGS, PSY_shared_-PGS). Models were adjusted for six principal components provided by the UK Biobank, as well as sex and year of birth. With previous studies demonstrating differences in educational attainment among participants who completed the UK Biobank Mental Health Questionnaire (MHQ) (37), we conducted a sensitivity analysis by performing linear regression on a subset of genotyped individuals (n=125,063) who took the 2016 MHQ.

#### C) Fluid Intelligence

The Fluid Intelligence measure in UKB (<u>ID: 20016</u>) consists of 13 reasoning questions, spanning verbal and numerical reasoning, logical puzzles, and pattern recognition. Participant score reflects the number of correct answers given; this was treated as a continuous variable. Linear regression models were performed again for each PGS to assess the relationship between genetic liability to psychotic disorders and fluid intelligence in 47,307 participants.

### Functional characterisation of SZ_specific_ and PSY_shared_ genetics

To assess the functional associations between genetic liability unique to schizophrenia and that which is shared with bipolar disorder, we leveraged the FUMA framework (v1.1.0) (41). The framework includes several analyses in MAGMA (v1.6) including gene-position and tissue expression analyses (42) (for full details see <u>Supplemental Note</u>).

### Mendelian Randomisation

Finally, to examine causal relationships between genetic liability to psychotic disorders with educational outcomes and IQ, two-way Mendelian randomisation (MR) was conducted. We selected genome-wide significant variants from summary-level GWAS data of schizophrenia (32), bipolar disorder (33), SZ_specific_, PSY_shared_, EA (43) and IQ (44). MR analysis was conducted using several complementary methods (see <u>Supplemental Note</u>), with the Inverse Variance Weighted (IVW) method serving as the primary test. Across all pairs, relationships were tested in both directions and Leave-One-Out (LOO) analyses were conducted to assess the impact of potentially influential variants. Diagnostic tests were employed to evaluate pleiotropy and heterogeneity. All MR analyses were performed using the TwoSampleMR package in R (45).

## Results

### 1. GWAS-by-subtraction

The results from the Genomic SEM model revealed significant relationships between schizophrenia, bipolar disorder, SZ_specific_, and PSY_shared_. The standardised loading from the PSYshared component on schizophrenia was 0.679 (SE = 0.024). The SZ_specific_ component, unique to schizophrenia, had a standardised effect of 0.734 (SE = 0.021). The analysis revealed that SZ_specific_ accounted for 53.9% of the total genetic variance in schizophrenia, with PSY_shared_ explaining the remaining 46.1% (Supplemental Table 2). GWAS-by-subtraction identified 63 independent genome-wide significant SNPs associated with SZ_specific_, distinct from 78 independent significant SNPs associated with PSY_shared_ (Figure 2). The effective sample size of the SZ_specific_ and PSY_shared_ GWAS were estimated to be 65,626 and 71,376, respectively.

**Figure 2:**
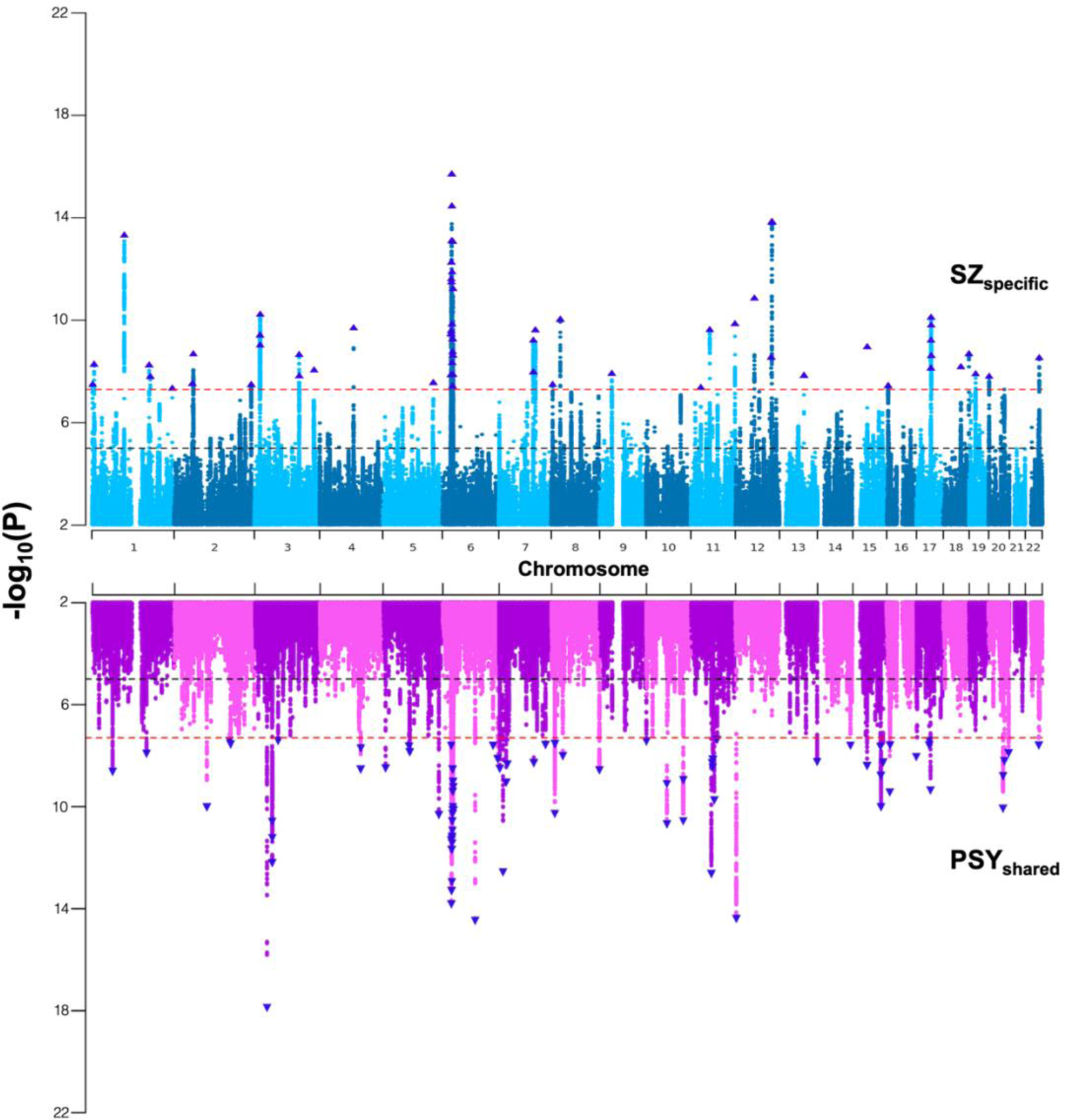
Miami Plot of GWAS for SZ_specific_ and PSY_shared_. The plot displays genome-wide association study (GWAS) results for SZ_specific_ (top, blue) and PSY_shared_ (bottom, purple) across chromosomes 1-22. Each point represents a single SNP, with the y-axis indicating the -log10(p-value) for association significance. The horizontal red dashed line indicates the genome-wide significance threshold (p < 5 × 10⁻⁸). Significant SNPs are highlighted above this threshold for SZ_specific_ and PSY_shared_ respectively.

### 2. Novel Genetic Associations

Of 63 significant SNPs independently associated with SZ_specific_, 16 have been reported in previous schizophrenia GWAS^41^, none had been reported in the most recent bipolar disorder GWAS^42^. Thirty-six SNPs had been reported in GWAS of other traits, including several which had been implicated in GWAS of brain structure and cognitive abilities. SNPs had also been reported in other GWAS including smoking, complement 4 levels and neuroticism (Supplemental Table 3). Notably, 22 of the 63 identified SNPs were novel associations, having not been previously reported for any trait in the GWAS catalog. In the PSY_shared_ GWAS, of the 78 independent significant SNPs, 15 were novel, whilst 47 had been previously associated with bipolar disorder. Five SNPs had been associated with educational attainment (EA), and 11 with schizophrenia (Supplemental Table 4).

### 3. Genetic Correlation analyses

Both schizophrenia and SZ_specific_ demonstrated a negative genetic correlation with IQ (SZ - IQ rg = -0.22, pFDR = 3.0E-22; SZsp - IQ rg = -0.24, pFDR = 2.8E-19), whilst the correlations between bipolar disorder and PSY_shared_ with IQ were much less pronounced (BD - IQ rg = -0.07, pFDR = 9.1E-04; PSY_shared_ - IQ rg = -0.07, pFDR = 2.3E-04). Schizophrenia exhibited minimal genetic correlation with EA (rg = 0.01, pFDR = 0.46), whereas the latent variables demonstrated divergent relationships (Figure 3); PSY_shared_ was positively correlated with EA (rg = 0.11, pFDR = 3.9E-09), whereas SZ_specific_ was negatively correlated (rg = -0.06, pFDR = 1.5E-02). All correlations were found to be significant, except for the correlation between schizophrenia and EA (Supplemental Table 5).

**Figure 3:**
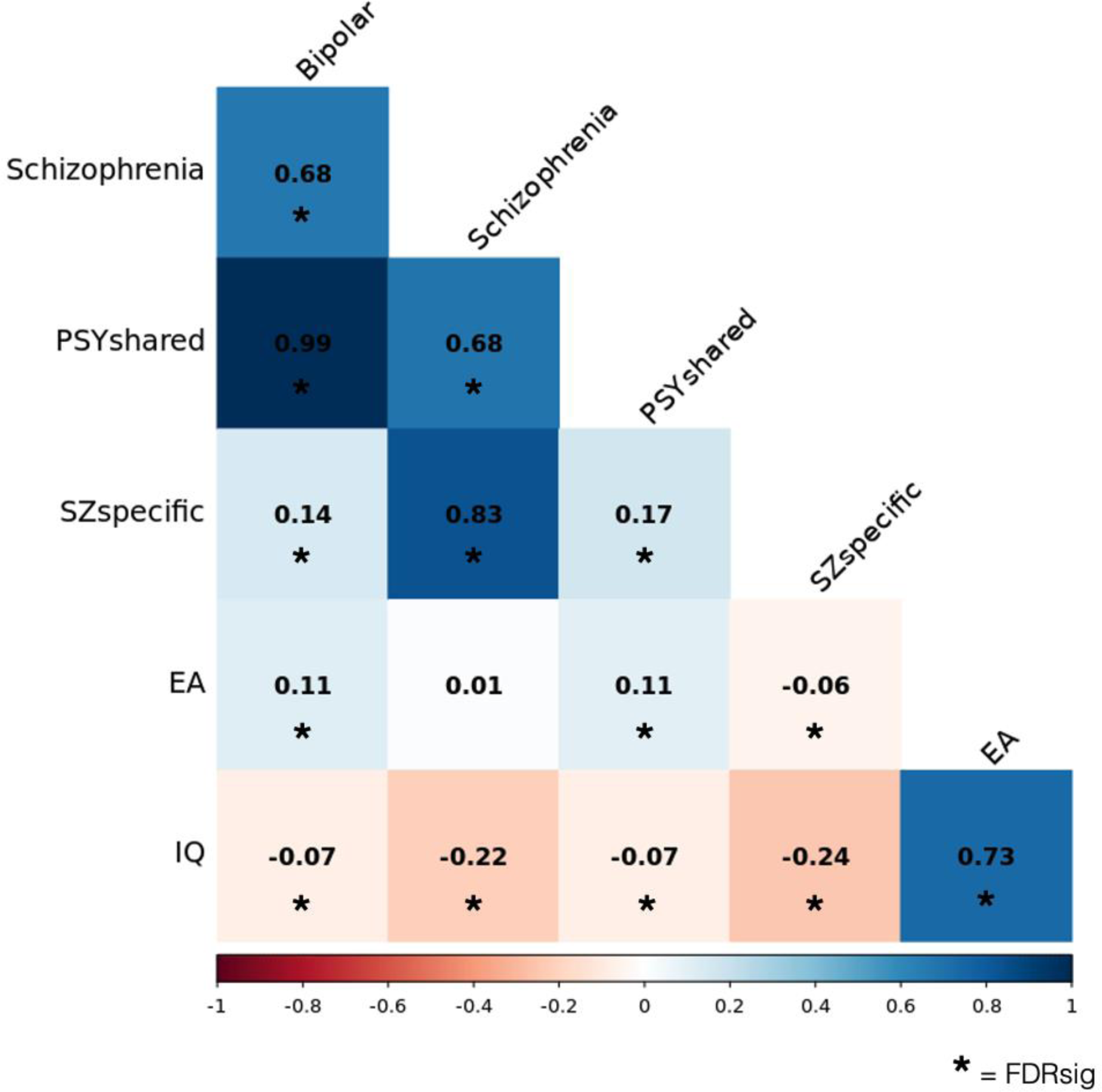
Genetic correlation matrix between bipolar disorder, schizophrenia, SZ_specific_, PSY_shared_, educational attainment (EA), and intelligence quotient (IQ). The heatmap displays genetic correlations, with the strength of the correlation (-1.0 – 1.0) indicated by the colour scale. Significant correlations (FDR-corrected) are marked with an asterisk (*).

### 4. Polygenic Score Analysis in the UK Biobank

#### 4.1 Educational Attainment

In our analysis of the UK Biobank, we examined the relationship between polygenic scores and educational attainment (EA) across two groups: the overall UK Biobank cohort and a subset who completed the Mental Health Questionnaire (MHQ). In the wider group, EA was calculated for 381,688 individuals, with a mean of 14.0 years of education (s.d. = 5.0). Among the MHQ respondents (n = 125,063), the average years in education was higher, at 15.7 years (s.d. = 4.6) (Supplemental Table 6).

The results of our linear regression analysis (Figure 4) revealed nuanced associations between polygenic scores for psychotic disorders, the latent variables and EA. In the broader UK Biobank cohort, the schizophrenia polygenic score (SZ-PGS) exhibited a small, non-significant negative association with EA (β = -0.002, p = 0.782), suggesting that genetic risk for schizophrenia does not substantially impact educational outcomes in this population. In contrast, the bipolar disorder polygenic score (BD-PGS) was significantly positively associated with EA (β = 0.14, p < 2e-16), indicating that genetic factors associated with bipolar disorder may be linked to higher educational attainment.

**Figure 4:**
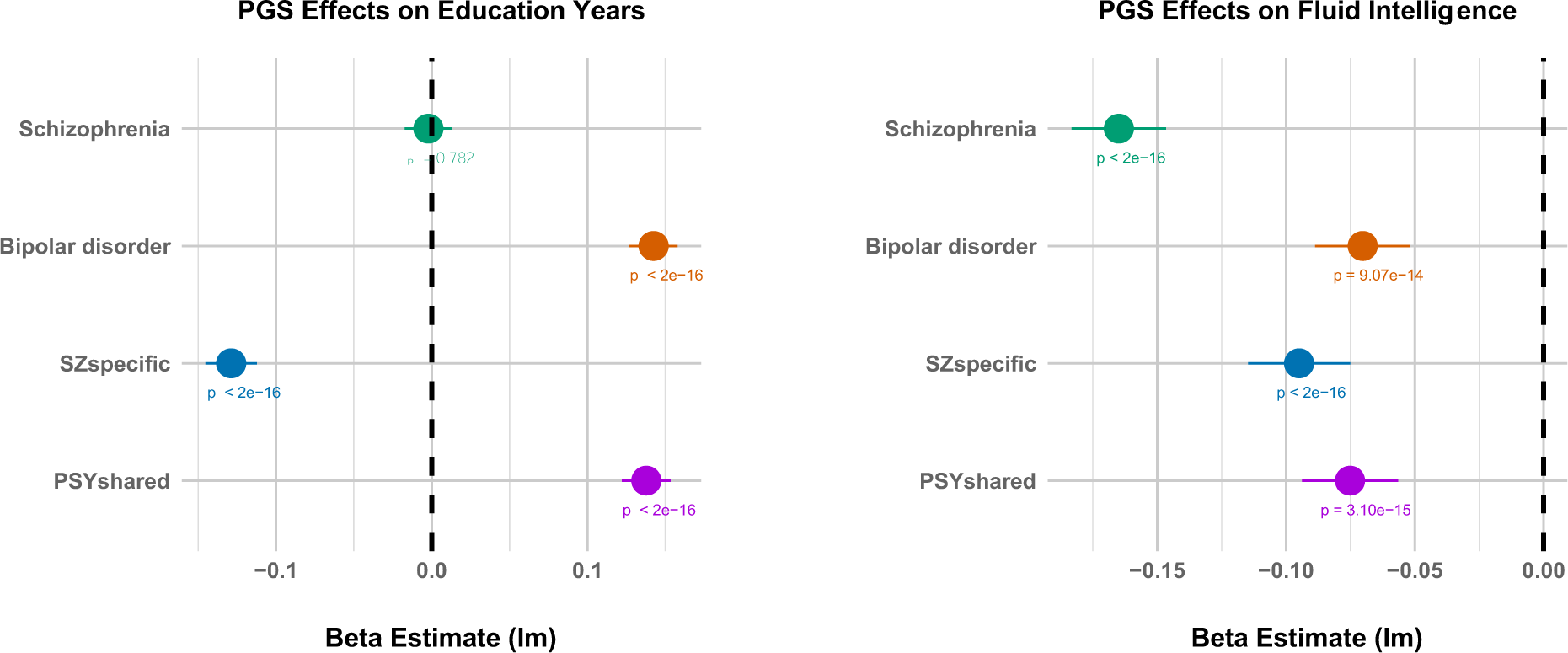
Impact of psychosis polygenic scores (PGS) on EA (educational attainment) and Fluid Intelligence in the UK Biobank. The plot on the left shows the effects of standardised PGS for schizophrenia, bipolar disorder, SZ_specific_ and PSY_shared_ on years of education. The plot to the right captures the effect of these PGS on fluid intelligence. Horizontal bars represent 95% confidence intervals. Beta estimates are derived from linear models (LM), with p-values detailed for significance.

The analysis of the latent variables revealed divergent effects on EA. SNPs unique to schizophrenia (SZ_specific_) had a significant negative association with EA (β = -0.13, p < 2e-16), reinforcing the notion that schizophrenia-specific genetic risk might hinder educational achievement. Conversely, SNPs shared between schizophrenia and bipolar disorder (PSY_shared_) were positively associated with EA (β = 0.14, p < 2e-16), suggesting that in contrast, the shared genetic component might be educationally advantageous.

When focusing on the MHQ subset, we observed that the general direction of these associations remained consistent with the overall cohort (Supplemental Table 7). However, the impact of SZ_specific_ was less pronounced (β = -0.08, p = 2.2e-09), and intriguingly, SZ-PGS showed a nominally significant positive association with EA (β = 0.03, p = 0.015).

#### 4.2 Fluid Intelligence

Fluid intelligence scores were available for 47,307 genotyped individuals within the UK biobank, with a mean score of 6.7 (s.d = 2.1). Our analysis revealed that all polygenic scores were significantly associated with reduced performance in the test (Figure 4), in order of effect: SZ-PGS (β = -0.16, p < 2e-16), SZ_specific_ (β = -0.09, p < 2e-16), PSY_shared_ (β = -0.08, p = 3.1e-15) and BD-PGS (β = -0.07, p = 9.1e-14). These findings suggest a spectrum of genetic risk impacting fluid intelligence from schizophrenia liability, through the latent variables to bipolar disorder.

### 5. Functional Characterisation of SZ_specific_ and PSY_shared_ SNPs

Five gene sets were significantly associated with the PSY_shared_ variable (Supplemental Table 8) predominantly related to synaptic signalling. No gene sets survived Bonferroni correction for the SZ_specific_ variable. Expression of SZ_specific_ and PSY_shared_ associated genes was highest in brain tissue compared to other general tissue types. PSY_shared_ gene expression was significantly enriched across all brain regions, with the highest relative levels of expression in cortical regions, notably the frontal cortex. Cortical expression was also high for SZ_specific_ genes, however, similar levels of gene expression were also seen in subcortical regions, such as the caudate and hippocampus (<u>Supplemental Note</u>).

### 6. Mendelian Randomisation (MR)

Across all exposure-outcome pairs, the genetic instruments demonstrated sufficiently high strength, with mean F-statistics ranging from 36.54 to 49.22. Measurement error levels were also low, indicated by high I² values (0.97 - 0.98). Full results for all MR analyses are in Supplemental Tables 9 - 12, Figures 5A and 5B.

**Figure 5:**
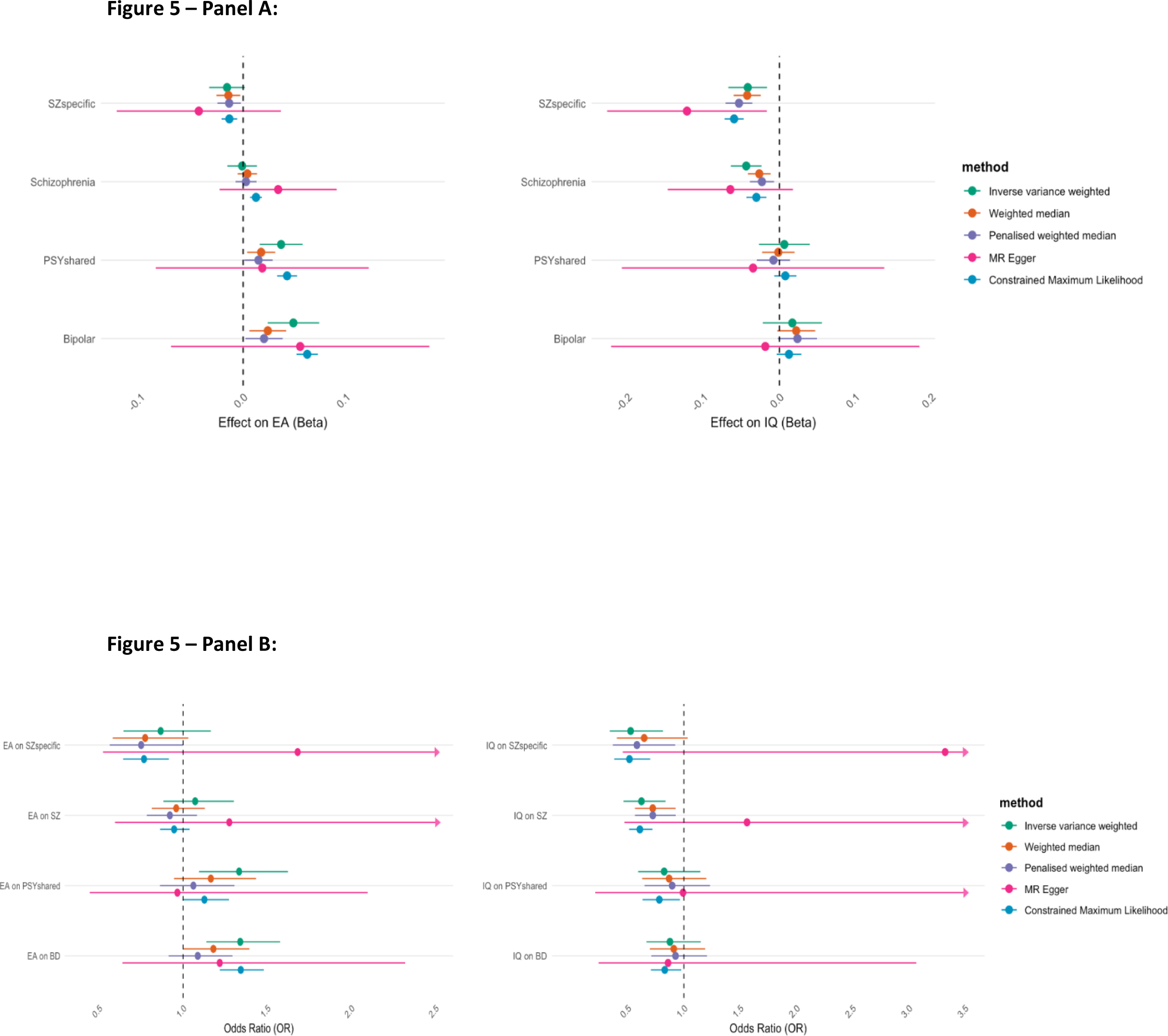
Panel A - Mendelian Randomisation (MR) estimates of the effects of SZ_specific_, PSY_shared_, schizophrenia, and bipolar disorder on educational attainment (EA) and intelligence quotient (IQ). The left panel shows the estimated effects (Beta) on EA, and the right panel shows the estimated effects on IQ (Beta). Several MR methods are used: Inverse variance weighted, weighted median, penalised weighted median, MR Egger, and constrained maximum likelihood. Horizontal bars represent 95% confidence intervals for each estimate, with the vertical dashed line indicating no effect (Beta = 0). **Panel B - Mendelian Randomisation (MR) estimates of the effects of educational attainment (EA) and intelligence quotient (IQ) on schizophrenia, bipolar disorder, SZ_specific_, and PSY_shared_.** The left panel presents the estimated odds ratios (OR) for the effect of EA on each psychosis component, while the right panel shows the OR for the effect of IQ. Multiple MR methods are applied: inverse variance weighted, weighted median, penalised weighted median, MR Egger, and constrained maximum likelihood. Horizontal bars indicate 95% confidence intervals, and the vertical dashed line represents an OR of 1, implying no effect.

#### 6.1 Psychotic Disorders and Educational Attainment

Our primary analysis using the inverse-variance weighted (IVW) method revealed no significant associations between schizophrenia and EA in either direction. In contrast, bipolar disorder exhibited a significant association with EA in both directions (IVW - BD on EA: β = 0.049, p = 0.0001; IVW - EA on BD: OR = 1.34, p = 0.0004). MR Egger estimates for both relationships were insignificant, suggesting the presence of genetic pleiotropy influencing these associations. The results of additional sensitivity analyses were consistent with the IVW estimates for the effect of bipolar disorder on EA. In the reverse direction (EA on bipolar disorder), only the contamination mixture likelihood (cML) estimate was significant, indicating potential variability in the robustness of these estimates.

The genetic components specific to schizophrenia (SZ_specific_) and those shared with bipolar disorder (PSY_shared_) showed divergent relationships with EA. Like bipolar disorder, PSY_shared_ was significantly positively associated with EA (IVW: β = 0.037, p = 0.0005). In the reverse direction, EA was also positively associated with PSY_shared_ (IVW: OR = 1.33, p = 0.0043). Sensitivity analyses for the effect of PSY_shared_ on EA were all significant except for the MR-Egger test. Conversely, for EA on PSY_shared_, only the IVW yielded a significant estimate. SZ_specific_ initially showed no significant association with EA (IVW: β = -0.016, p = 0.08). However, after removing a highly influential outlier variant (rs4950119) identified through leave-one-out analysis, this negative relationship became significant (IVW: β = -0.021, p = 0.01). This finding was supported by all sensitivity analyses, except for the MR Egger test, which remained non-significant. No significant association was found for EA on SZ_specific_.

#### 6.2 Psychotic Disorders and IQ

A significant negative association was found between schizophrenia and IQ (IVW - SZ on IQ: β = -0.04, p < 0.0001; IVW - IQ on SZ: OR = 0.62, p = 0.002), suggesting that genetic liability to schizophrenia is linked to lower cognitive performance. Sensitivity analyses largely supported these findings, with all methods except MR-Egger showing significant results. In contrast, no significant associations were found between bipolar disorder or PSY_shared_, with IQ in either direction. SZ_specific_ had a significant negative impact on IQ in the forward direction (IVW: β = -0.04, p < 0.001). All sensitivity tests were consistent in both direction and significance. IQ was also significantly associated with SZ_specific_ in the reverse direction with (IVW: OR = 0.53, p = 0.004). Sensitivity analyses further supported this finding, with significant results in the cML and penalised weighted median tests, although MR-Egger and the weighted median were non-significant.

#### 6.3 Pleiotropy and Heterogeneity

Given the non-significant results from most MR Egger regressions, we tested the intercepts to assess for the presence of directional pleiotropy across all exposure-outcome pairs. These tests were non-significant, indicating that whilst present, the observed pleiotropy is unlikely to bias our results. Cochran’s Q values were, however, large and highly significant across all comparisons. These findings suggest substantial heterogeneity among the genetic instruments, and highlight that multiple pathways may exist between the exposures and outcomes (46), contributing to the observed variability.

## Discussion

### 1. Summary of findings

Using a GWAS-by-subtraction of psychotic disorders we identified 63 independent loci unique to schizophrenia (SZ_specific_) and 78 loci that are shared with bipolar disorder (PSY_shared_). Our analyses reveal divergent relationships between components of schizophrenia liability and educational attainment (EA). SZ_specific_ was negatively associated with EA, whereas PSY_shared_ was positively associated, providing strong evidence to support the hypothesis that the positive association between schizophrenia genetic risk and EA appears driven by shared liability with bipolar disorder. Additionally, we highlight that genetic risk unique to schizophrenia may be more deleterious to cognitive ability, supporting the notion of a neurodevelopmental pathway to psychotic disorders.

### 2. Findings in context

Numerous studies have described positive associations between genetic risk for schizophrenia and EA (16, 20, 21), prompting several groups to examine this paradox. In the UK Biobank, Escott-Price et al. found SZ-PGS to be negatively associated with both ordinally ranked educational attainment and when qualifications were dichotomised into ‘Academic Qualifications’ versus others (47). They suggested that earlier positive associations resulted from smaller sample sizes (16) and the comparison of dichotomised higher (University and College) versus lower educational qualifications. They hypothesised that socio-demographic factors, rather than cognitive skills, might drive educational attainment in the UK. This was not supported, however, by analysis of IQ-PGS, which showed a strong positive linear relationship across all attainment groups. Several multivariate Mendelian randomisation (MR) studies have since explored other potential confounders to this relationship. They have revealed that when controlling for income and cognition, the small positive effect of schizophrenia on EA remains, however, the association between bipolar disorder liability and EA has been shown to be stronger (27).

Current psychiatric classifications dichotomises schizophrenia and bipolar disorder without clear evidence for neurobiological segregation (28). The disorders have a high degree of genetic correlation (rg = 0.7) (48) and share significant comorbidity due to overlapping genetic risk (11). Several symptoms characteristic of each diagnosis are seen in the other; psychosis is common in bipolar disorder (49), whilst depression is frequently reported in schizophrenia (50). One significant point of phenotypic separation, however, is cognition (4). In contrast to schizophrenia, a bipolar diagnosis may be enriched in those at the extreme high end of the intelligence spectrum (51). This epidemiological evidence, in combination with overlapping genetic risk, has hinted at the potentially confounding impact of bipolar disorder liability on the relationship between schizophrenia liability and EA. Adams demonstrated that while genetic liability to more years in education seemed to increase schizophrenia risk, the effect disappeared when bipolar disorder liability was included in the model (29). These findings suggested that the association between higher education and increased schizophrenia risk may be a product of the pleiotropic effects of bipolar disorder liability on both schizophrenia and education. Attempts to delineate the relative contribution of genetic risk that is unique to schizophrenia on EA have supported this conclusion. Using a genome-wide inferred statistics approach (GWIS), Bansal et al., reported a negative genetic correlation between EA and schizophrenia risk that was separated from risk shared with bipolar disorder (52), which contrasted to a positive correlation between EA and schizophrenia overall. Genomic SEM analyses have provided further support. Regressing the genetic component of EA onto the genetic components of bipolar disorder and schizophrenia the seminal Grotzinger et al., paper demonstrated a larger association with EA for bipolar disorder than schizophrenia (30).

Building on these foundations, we have provided further evidence to support the hypothesis that positive associations between schizophrenia and EA are a product of the genetic risk that schizophrenia shares with bipolar disorder. We demonstrated divergent associations between polygenic scores for psychotic disorders, their component parts, and EA. Consistent with the Escott-Price et al., analysis (47), SZ-PGS showed a small, non-significant negative association with EA in the overall UK Biobank cohort, while in the MHQ sample, the relationship was positive, possibly as a result of underlying attainment disparities between these cohorts (37). In contrast, BD-PGS consistently demonstrated significant positive associations with EA, reinforcing the idea that bipolar disorder’s genetic architecture may contribute to enhanced educational outcomes. When dissecting schizophrenia risk further, SNPs specific to schizophrenia (SZ_specific_) were significantly negatively associated with EA, whereas schizophrenia SNPs shared with bipolar disorder (PSY_shared_) were significantly positively associated with EA. This suggests that genetic factors unique to schizophrenia may be more deleterious to EA, highlighting a distinction between the educational impacts of SZspecific and PSYshared genetic components. Mendelian Randomization (MR) analyses offered preliminary support for our findings, where we demonstrated a bidirectional positive association between PSY_shared_ and EA and a predominately negative relationship between SZ_specific_ and EA.

Our analyses revealed significant associations between polygenic scores for psychotic disorders and fluid intelligence in the UK Biobank. All polygenic scores were significantly associated with reduced fluid intelligence scores, with SZ-PGS having the most pronounced effect, followed by SZ_specific_-PGS, PSY_shared_-PGS and BD-PGS, each contributing to lower performance on fluid intelligence tests. MR findings supported these results, but also demonstrated a continuum of effect with significant negative bidirectional relationships between IQ and both SZ_specific_ and overall schizophrenia risk, in order of effect size. Bipolar disorder and PSY_shared_ were not significantly associated with IQ. These results align with our genetic correlation analyses, demonstrating negative correlations between IQ and both schizophrenia and SZ_specific_, with less pronounced negative correlations with bipolar disorder and PSY_shared_. A continuum of cognitive impairment across psychotic disorder liability has also previously been supported by MR results from Ohi et al., (15) who leveraged results from a unique GWAS, directly comparing individuals with schizophrenia and bipolar disorder (53). They found bidirectional relationships between intelligence and schizophrenia risk, with lower intelligence unidirectionally associated with schizophrenia specific risk. In keeping with our findings, intelligence was unrelated to bipolar risk, however, the authors did find an association between intelligence and shared risk between schizophrenia and bipolar disorder, which was not repeated in our study.

### 3. Dissecting clinical heterogeneity in psychotic disorders with genetics

While reaffirming the lack of clear biological divergence between schizophrenia and bipolar disorder, our study highlights the power of genetics to unravel the clinical heterogeneity within these diagnoses. Polygenic scores (PGS) for psychotic disorders have previously demonstrated diverse associations with different symptom clusters. For instance, SZ-PGS correlates with negative and disorganised symptoms but not with positive symptoms, which are more commonly shared with bipolar disorder (54). The genetic complexity extends to cognitive symptoms as well. Although SZ-PGS is linked to poorer cognitive performance in healthy controls, findings in individuals with schizophrenia have been inconsistent (54, 55). In contrast, rare coding and copy-number variants (CNVs) associated with schizophrenia consistently worsen cognitive function in patients (56). These CNVs (57) are also enriched in individuals with autism and intellectual disability (58), but have been less strongly associated with bipolar disorder (59).

These findings have prompted the conceptualisation of psychotic disorders as existing on a cognitive continuum with neurodevelopmental disorders, with an increasing rare genetic burden towards the more severe end of the spectrum (19). Our study provides preliminary evidence that common variants unique to schizophrenia (SZ_specific_) may also be more neurodevelopmentally deleterious than those shared with bipolar disorder (PSY_shared_). This aligns with the Murray et al. hypothesis that schizophrenia comprises both a neurodevelopmental disorder characterised by cognitive impairment and negative symptoms, and a second disorder more aetiologically similar to bipolar disorder (60). This neurodevelopmental component of schizophrenia was hypothesised to be linked to genetic or early environmental damage to the hippocampus. Whilst PSY_shared_ genes are most highly expressed in cortical regions, SZ_specific_ genes are also expressed at similar levels in subcortical regions, such as the caudate and hippocampus. The hippocampus is both central to cognitive function in health and has been strongly implicated in the cognitive symptoms of schizophrenia (4). Increasing evidence suggests that an excitatory/inhibitory imbalance in the hippocampus may be a key driver of several symptoms in schizophrenia (61), particularly in individuals with 22q11.2 deletions (62), the strongest known genetic risk factor for the disorder.

There are many pathways to psychosis, likely reflecting diverse underlying aetiologies. It is increasingly evident that heterogeneity, both clinical and genetic, is the rule rather than the exception, and that current diagnostic criteria often fail to capture the full biological complexity of psychotic disorders. Indeed, initial modelling of psychosis biotypes by the BSNIP consortium revealed that psychotic diagnoses primarily reflected a continuum of cognitive ability, rather than distinct neurobiological entities (63). However, further modelling demonstrated that these biotypes possess fundamentally opposing neurophysiological signatures, indicating that similar symptoms across individuals may arise from different underlying disease processes.

In this context, our work contributes to a growing body of literature suggesting that the statistical dissection of genetic risk for psychotic disorders offers deeper insights into underlying disease mechanisms than disorder-level PGS alone. Early efforts to develop neurotransmitter pathway-specific PGS are beginning to support this approach. Although based on small samples, glutamatergic PGS have been shown to more accurately index deficits in cognitive control in schizophrenia, while dopaminergic PGS are more closely associated with impairments in global functioning (64). It is notable that the majority of cases of treatment resistant schizophrenia have glutamatergic rather than dopaminergic dysfunction (65), and that these cases have been associated with both neurodevelopmental cognitive impairment (66) and an increased burden of schizophrenia-associated CNVs (67).

## Limitations

The interpretation of this study’s findings must be contextualised within its limitations. Using Genomic SEM, we have modelled latent variables and constructed a simplified path diagram to represent component parts of schizophrenia genetic risk. Though Genomic SEM is widely used, our model, like any, cannot suppose exact concordance with nature. The instruments used in the GWAS-by-subtraction each also have inherent limitations. Case-control status in the GWAS of psychiatric disorders are based on categorical criteria, thus suffering from diagnostic heterogeneity. These GWAS often consist of meta-analysed cohorts, each with their respective ascertainment biases. A notable example is the CLOZ-UK cohort within the schizophrenia GWAS, which consists of clinically ascertained individuals with more severe, treatment-resistant forms of schizophrenia (68). The cognitive ability of these participants is likely to differ from cohorts that followed individual consenting procedures, potentially introducing bias when comparing genetic effects across studies. Additionally, EA and IQ GWAS are often heavily confounded by environmental factors and ascertainment biases, which might affect the accuracy of polygenic scores used in this analysis. Moreover, due to the assumptions behind Genomic SEM, GWAS were limited to individuals of European ancestry, thus reducing the generalisability of findings.

The UK Biobank, though widely used in genetic research, also has notable limitations. Participants are generally healthier, older, more educated, and from higher socioeconomic backgrounds compared to the general UK population - when considering associations between these variables and individuals with severe mental illness - it is clear that the UK biobank would not be in keeping with clinical samples (67). Studying EA in the UK Biobank is also context specific. Given the focus on the UK educational system, as well as variations in educational policy across the age range of participants, the generalisability of findings are of course limited to the setting in which they are examined. Another possible limitation in the analysis concerns MR. Genetic variants can be pleiotropic and influence multiple traits; this can bias estimates if they affect the outcome through pathways other than the exposure of interest. We have used pleiotropy robust methods in this study to counter this where possible.

## Conclusion

Our study highlights the value of dissecting genetic risk in neuropsychiatric disorders to better understand their complex phenotypes. We have demonstrated that GWAS-by-subtraction can reveal complex associations between components of schizophrenia genetic risk with cognitive and educational outcomes. More broadly we provide provisional evidence to suggest that schizophrenia genetic liability may be better understood through two components: one overlapping with bipolar disorder, and another akin to neurodevelopmental disorders. Future studies should explore these components in diverse and deeply phenotyped cohorts to further elucidate the causal roots of these conditions.

## Data Availability

All data produced in the present study are available upon reasonable request to the authors.

## Acknowledgements

This research is funded by the National Institute for Health and Care Research (NIHR) Maudsley Biomedical Research Centre (BRC). The views expressed are those of the authors and not necessarily those of the NIHR or the Department of Health and Social Care. CJW is supported by an NIHR Academic Clinical Fellowship (ACF-2022-17-009). LG is funded by the King’s College London DRIVE-Health Centre for Doctoral Training and the Perron Institute for Neurological and Translational Science.

