## Supplemental Note for "Splitting Schizophrenia: Divergent Cognitive and Educational Outcomes Revealed by Genomic Structural Equation Modelling"

#### GWAS summary statistic preparation

Publicly available GWAS summary statistics for schizophrenia (1), bipolar disorder (2), IQ (3), and educational attainment (EA) (4) were used in this study. Due to the sensitivity of Genomic SEM and Linkage Disequilibrium Score Regression (LDSC) methods to ancestral variation, only summary statistics derived from samples of European ancestry were selected for analysis. Detailed information on each cohort can be found in Supplementary table 1.

#### Genomic SEM parameters

Each of the above summary statistics were prepared in turn for use in Genomic SEM (5). Mean allele frequency (MAF) scores were obtained from the 1000 Genomes Project (6), with SNPs filtered out if MAF was <0.01. SNPs were also removed if the imputation quality (INFO) score was <0.6. The effective sample size (NEFF), describing the sample size of a balanced selection of cases and controls with the equivalent statistical power, was obtained from the GWAS summary statistics for each binary trait (schizophrenia = 117,498, bipolar = 101,962).

#### GWAS-by-subtraction method

Following the method proposed by Demange et al 2021 (7), we first regressed GWAS summary statistics for schizophrenia and bipolar disorder onto two latent variables, named '$SZ_{specific}$' and '$PSY_{shared}$'. We specified that all the covariance in schizophrenia and bipolar disorder would be captured by these latent variables; their variance was set to 1, and their covariance to 0. Following Demange's model, the error on the two manifest variables was set to 0. After fitting the model, our latent variables were then regressed onto the SNPs contained within each GWAS. The model (Figure 1) resulted in one path representing the genetic effects shared between schizophrenia and bipolar disorder ($PSY_{shared}$), while the other path measured the SNP effects unique to schizophrenia ($SZ_{specific}$), independent of shared effects. The NEFF for

$PSY_{shared}$ and $SZ_{specific}$ was calculated as suggested in Demange et al 2021. Since the GWAS-by-subtraction model is fully saturated, no fit statistics were produced during the analysis.

**Genetic correlation analyses**

The LDSC function in the Genomic SEM package (5) was used to calculate the genetic correlation between schizophrenia, bipolar disorder, $SZ_{specific}$, $PSY_{shared}$, EA and IQ. Summary statistics were converted to a form usable in LDSC, aligning alleles to the HapMap3 reference set and imposing INFO and MAF filters of >0.9 and >0.01 respectively. We conducted multivariable LDSC using sample prevalences of 0.5 for SZ, BD, $SZ_{specific}$ and $PSY_{shared}$ as recommended when using NEFF (7). Population prevalence for bipolar disorder and schizophrenia was defined as 2% and 0.7% respectively as specified in the original disorder GWAS (1, 2). LDSC then generated a genetic covariance matrix and genetic correlation for each trait pair. P-values were calculated for each pair, testing the hypothesis that the estimates are different from zero. A Benjamini-Hochberg False Discovery Rate (FDR) was applied to these p-values (q-value = 0.05) to mitigate for multiple testing bias.

**Identifying significant SNPs and novel associations**

Using PLINK v1.9 (8) we identified independently associated SNPs for $SZ_{specific}$ and $PSY_{shared}$. In concordance with the original GWAS-by-subtraction paper, we defined independent SNPs as either falling outside a 250 kb window, or being within that window, but with a linkage disequilibrium (LD) r2 < 0.1. Data on LD was obtained from the 1000 Genomes Project (EUR N = 503) (6). Significance was defined using the genome wide significance threshold of $p < 5 \times 10^{-8}$. Searching the GWAS Catalog (9), each significant SNP was compared to previously published GWAS of other traits, including schizophrenia and bipolar disorder. SNPs were considered novel if they had not previously been reported in the GWAS Catalog.

**Polygenic score creation in GenoPred and Quality Control**

Polygenic scores were created based on summary data from schizophrenia and bipolar disorder GWAS and the GWAS of our latent variables, $SZ_{specific}$ and $PSY_{shared}$. PGS were calculated using MegaPRS (10), leveraging the GenoPred pipeline (11). GenoPred infers genetic ancestry by comparing target data with both the 1000 Genomes Phase 3 (1KG) and Human Genome Diversity Project (HGDP) samples (12), restricted to HapMap3 variants only. Ancestry was inferred for 487,409 individuals in the UK Biobank. European ancestry was assigned to 456,793 individuals for whom polygenic scores were calculated. After quality control procedures removing individuals with high missingness (>2%), abnormal sex checks and QA errors, 453,541 individuals remained. Lastly, we accounted for individuals with high relatedness (kinship >0.044) to other participants, retaining one individual from each related pair. The resulting cohort contained 381,688 participants for whom sex and year of birth data was available.

**Educational attainment (EA) in the UK Biobank**

Educational attainment (EA) (ID: 6138) was assessed via touchscreen questionnaire, with participants reporting their highest level of qualification from the following options: "Other professional qualifications", "NVQ or HND or HNC or equivalent", "CSEs or equivalent", "O levels/GCSEs or equivalent", "A levels/AS levels or equivalent", and "College or University degree". The highest level reported by participants across four possible visits was taken, and replicating methods implemented in recent large studies (13), we derived an Education Years estimate. The Education Years estimate is based on the average number of years in education associated with each qualification. Linear regression models were used to examine the relationship between Education Years and polygenic scores (SZ-PGS, BD-PGS, $SZ_{specific}$-PGS, $PSY_{shared}$-PGS). Models were adjusted for six principal components provided by the UK Biobank, as well as sex and year of birth. Given the potential for societal influences on education attainment, including changes to the amount of mandatory education required in the UK, we assessed both age and year of birth as covariates in our regressions. With both variables highly collinear, we used year of birth, given its likely better indexing of educational context. With previous studies demonstrating differences in educational attainment among participants who completed the UK Biobank Mental Health Questionnaire (MHQ) (14), we conducted a sensitivity analysis by performing linear regression on a subset of genotyped individuals (n=125,063) who took the 2016 MHQ.

**Functional characterisation of $SZ_{specific}$ and $PSY_{shared}$ genetics**

To assess the functional associations between genetic liability unique to schizophrenia and that which is shared with bipolar disorder, we leveraged the FUMA framework (v1.1.0) (15), which includes several analyses in MAGMA (v1.6) (16). Gene-based analyses were conducted to positionally map SNPs to genes within a max distance of 10kb. Gene-set analyses were then conducted based on "curated gene sets" and "GO terms" from the Molecular Signatures Database v7.0 (17), assessing the relative strength of associations between gene sets and each latent variable GWAS. Finally, gene-property analysis was performed to evaluate the relationship between genetic associations and gene expression profiles. This was conducted using RNAseq data from the GTEx v8 database (18), covering 30 general tissue types and 53 specific tissue types. Gene sets and tissues were considered significant if the Bonferroni-corrected p-value was <0.05 within each tissue analysis.

**Mendelian Randomisation**

**A) Genetic instruments**

To examine causal relationships between genetic liability to psychotic disorders with cognitive and educational outcomes, two-way Mendelian randomisation (MR) was conducted. We used variants from summary-level GWAS data of schizophrenia (1), bipolar disorder (2), $SZ_{specific}$, $PSY_{shared}$, EA (4) and IQ (3) as instrumental variables. Variables were retained if they were statistically independent based on a clumping threshold of 10,000 kb and R2 < 0.001. Data on LD was obtained from the 1000 Genomes Project (EUR N = 503) (6). Variants were included as instruments if they reached the genome wide significance threshold of $p < 5e-8$. For each exposure-outcome pair, SNPs were first harmonised, before removing ambiguous and palindromic variants. To assess the strength of our genetic instruments and the potential for measurement error, we calculated F- and I-squared statistics respectively.

#### B) MR Analyses

MR analysis was conducted using several complementary methods. The Inverse Variance Weighted (IVW) method served as the primary test, providing a weighted average of the ratio estimates. However, this method assumes no horizontal pleiotropy influencing the MR associations (19). To address potential pleiotropy, sensitivity analyses were conducted if the IVW estimate was significant. The MR-Egger (20) is the most parsimonious approach, allowing all variants to have pleiotropic effects, while the weighted-median estimator can provide reliable estimates as long as at least half of the variants have no pleiotropic effects (21). We also applied the Penalised Weighted Median method, which penalises the effects of heterogeneous instruments. Lastly, we applied the Constrained Maximum Likelihood (cML) method, which is robust provided that at least a plurality of instruments are valid (22). Across all pairs, Leave-One-Out (LOO) analysis was performed to assess the influence of individual variants and visually inspect for outliers through forest plots. Diagnostic tests were also employed to evaluate pleiotropy and heterogeneity: the MR-Egger intercept test, where a p-value $< 0.05$ indicates significant directional pleiotropy, and Cochran's Q test for heterogeneity, with $p < 0.05$ suggesting the presence of invalid instruments. We considered evidence of causality if the IVW estimate was significant and directionally consistent with all sensitivity analyses, without any influential variants in LOO analysis, no significant MR-Egger intercept, and no significant heterogeneity. All analyses were performed using the TwoSampleMR package in R (23).

### Supplementary Figures

**Figure 1:** QQ plots for GWAS of psychosis components

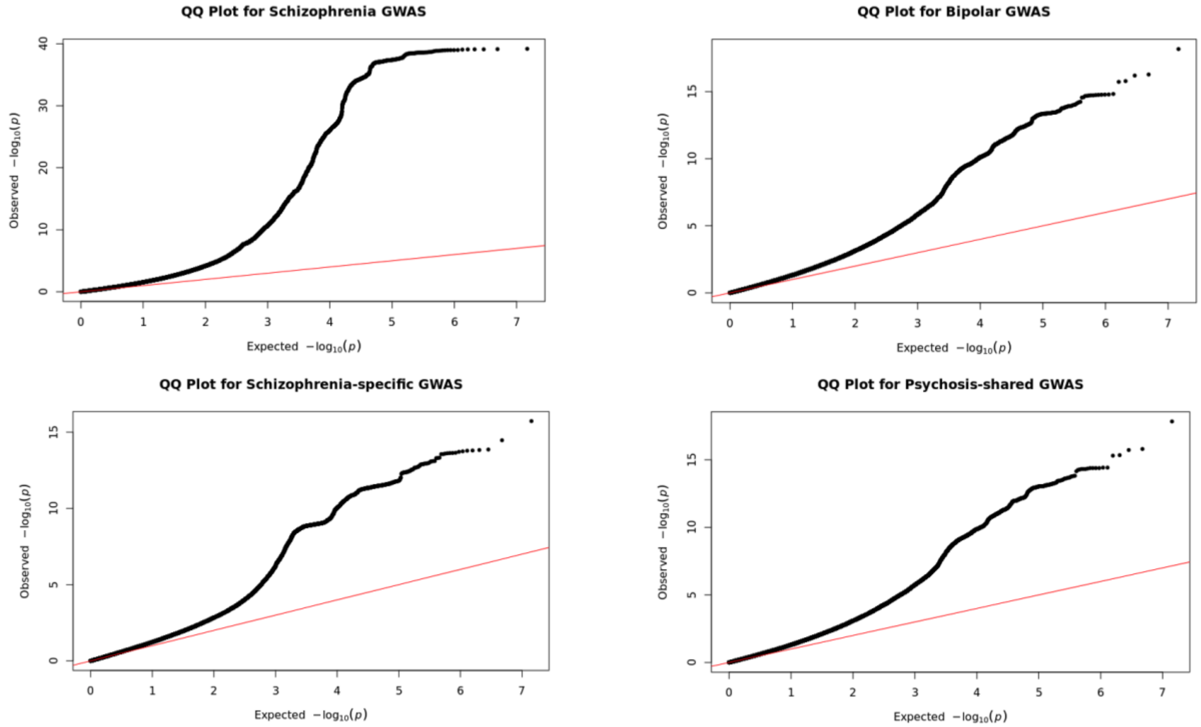


**Figure 2:** Distribution of psychosis PGS within the UK Biobank sample

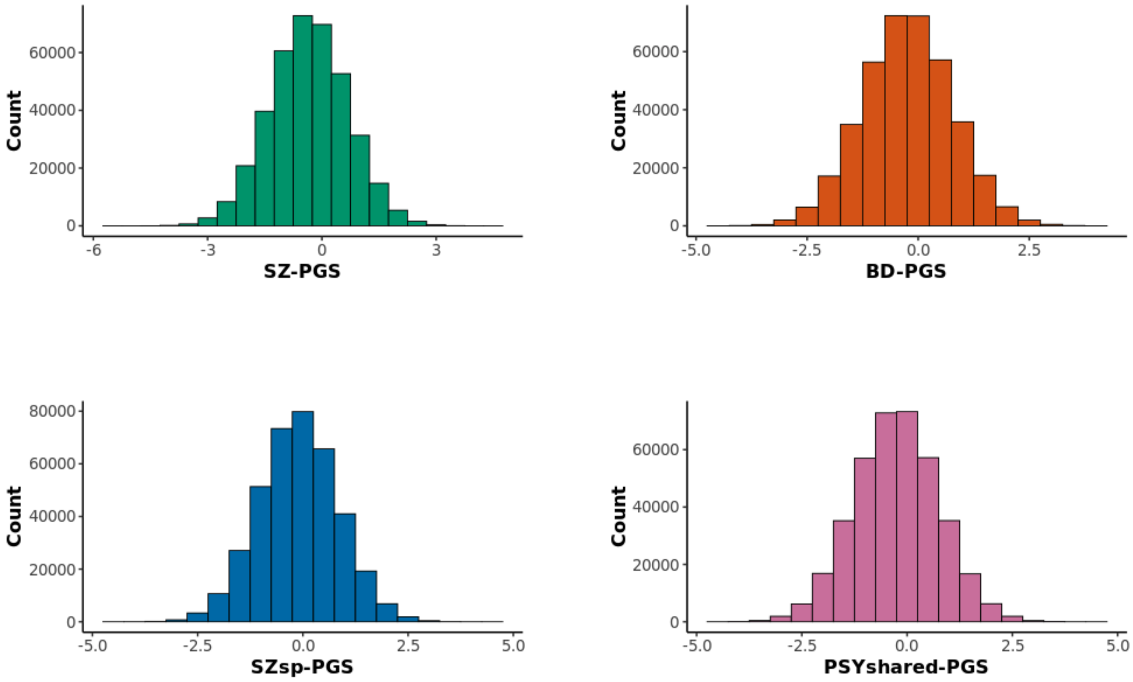

**Figure 3:** Distribution of Education Years (Whole UKB Cohort)

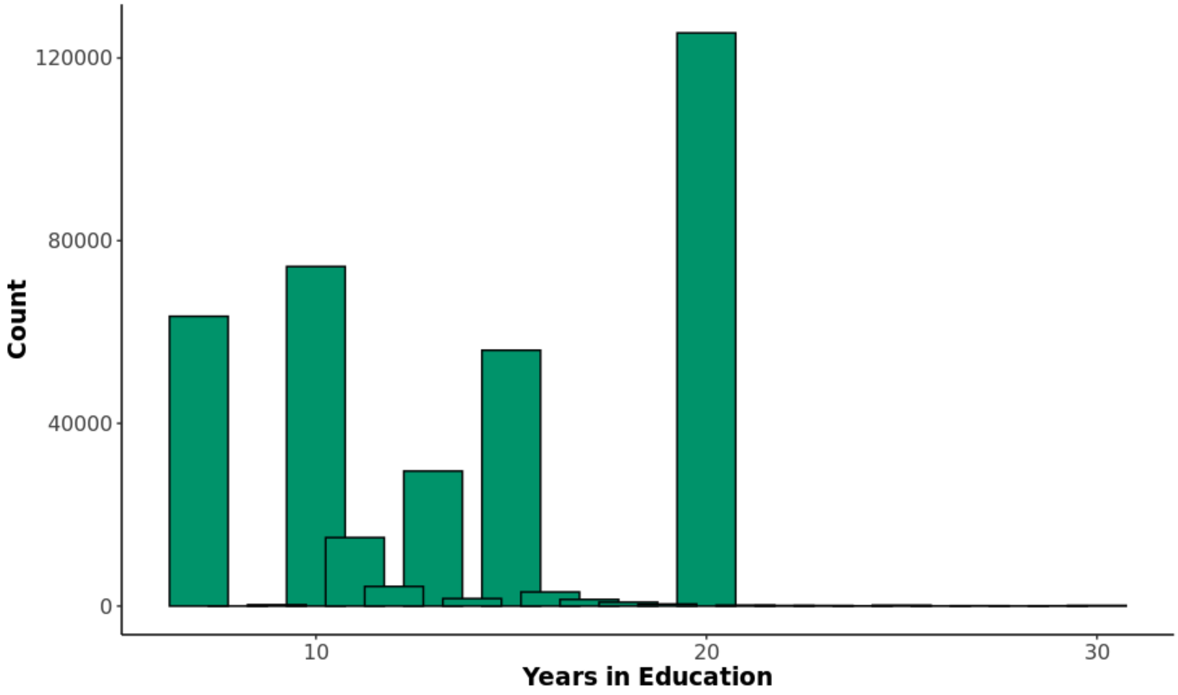


**Figure 4:** Distribution of Education Years (MHQ Subset)

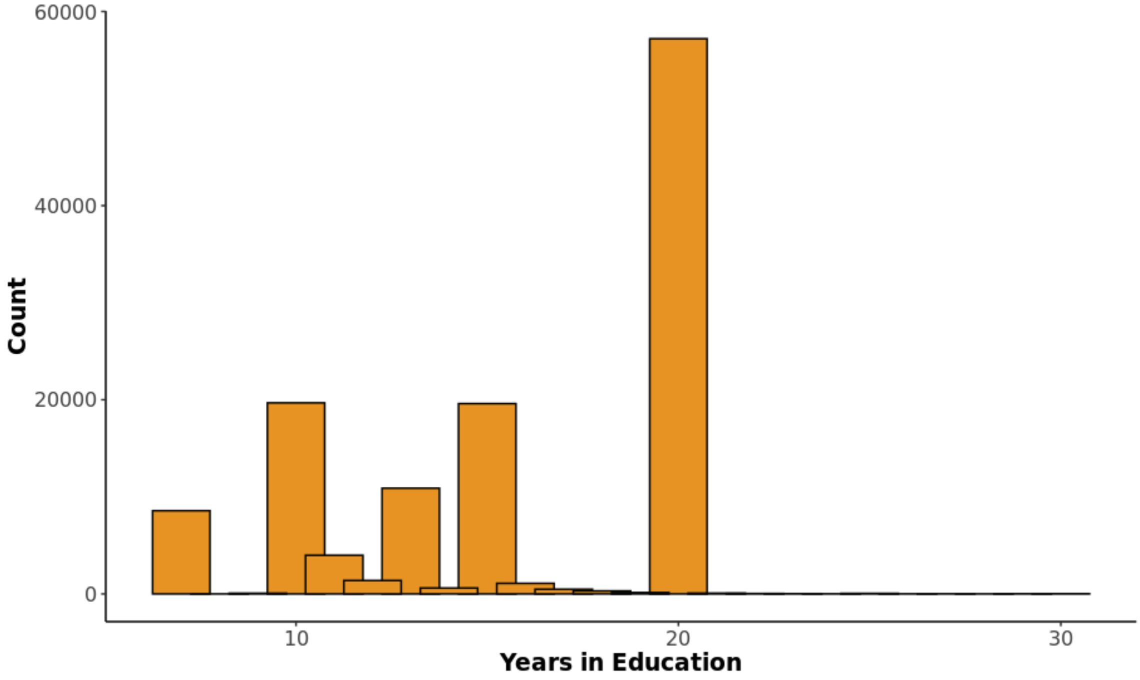

**Figure 5:** Forest Plot of PGS Effects on Education Years - Whole UKB Cohort vs UKB Sample

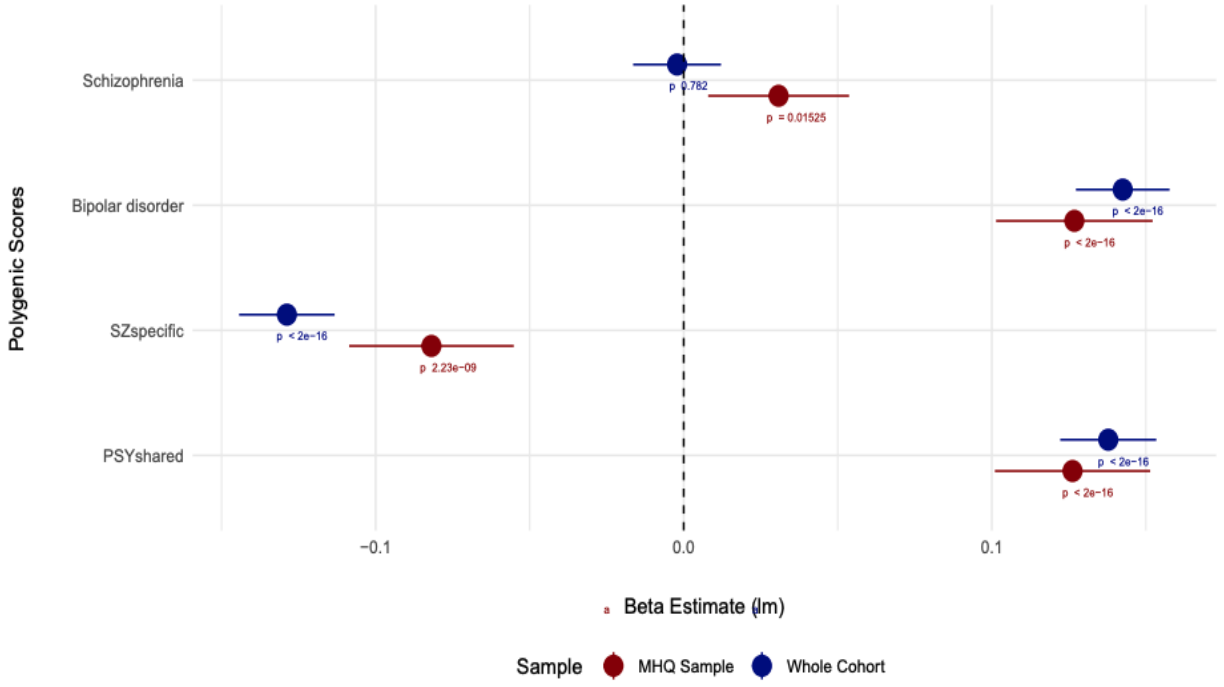


**Figure 6:** Distribution of Fluid Intelligence score results

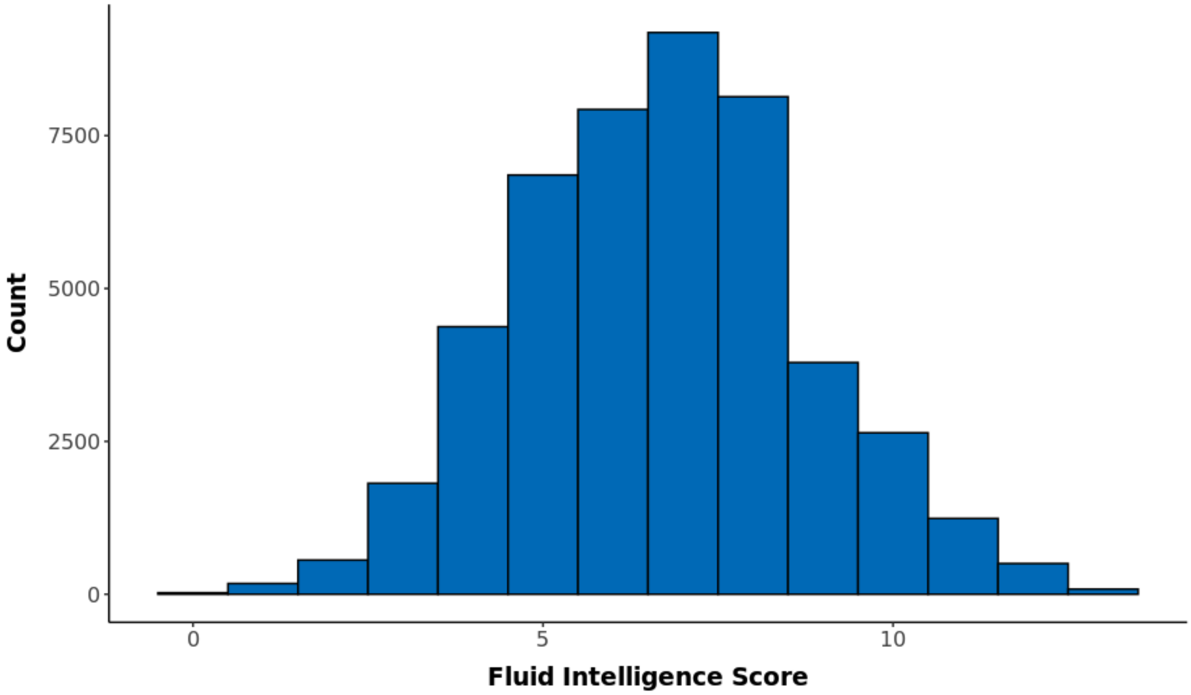

**Figure 7:** Top 100 Genes for SZspecific from gene-based analyses

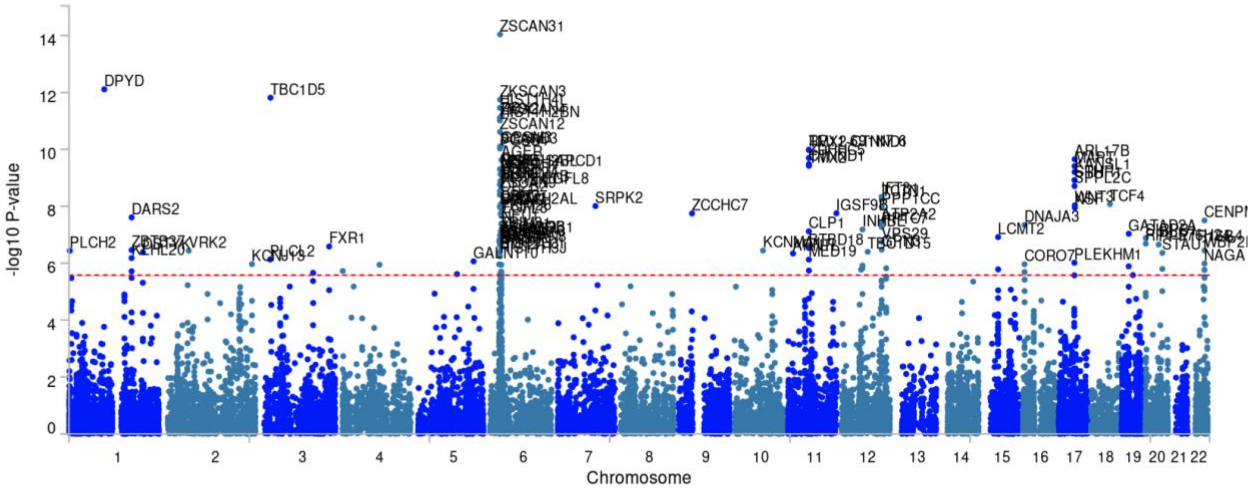


**Figure 8:** MAGMA Tissue Expression Analysis - SZspecific (GTEx v8 30 general tissue types)

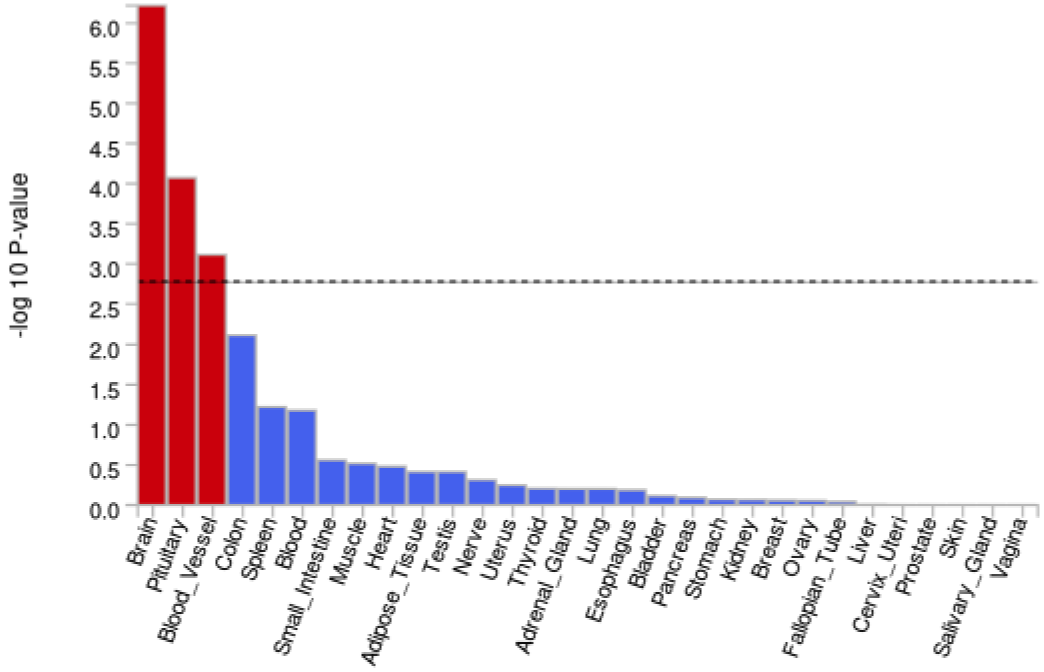

**Figure 9:** MAGMA Tissue Expression Analysis - SZspecific (GTEx v8 53 tissue types)

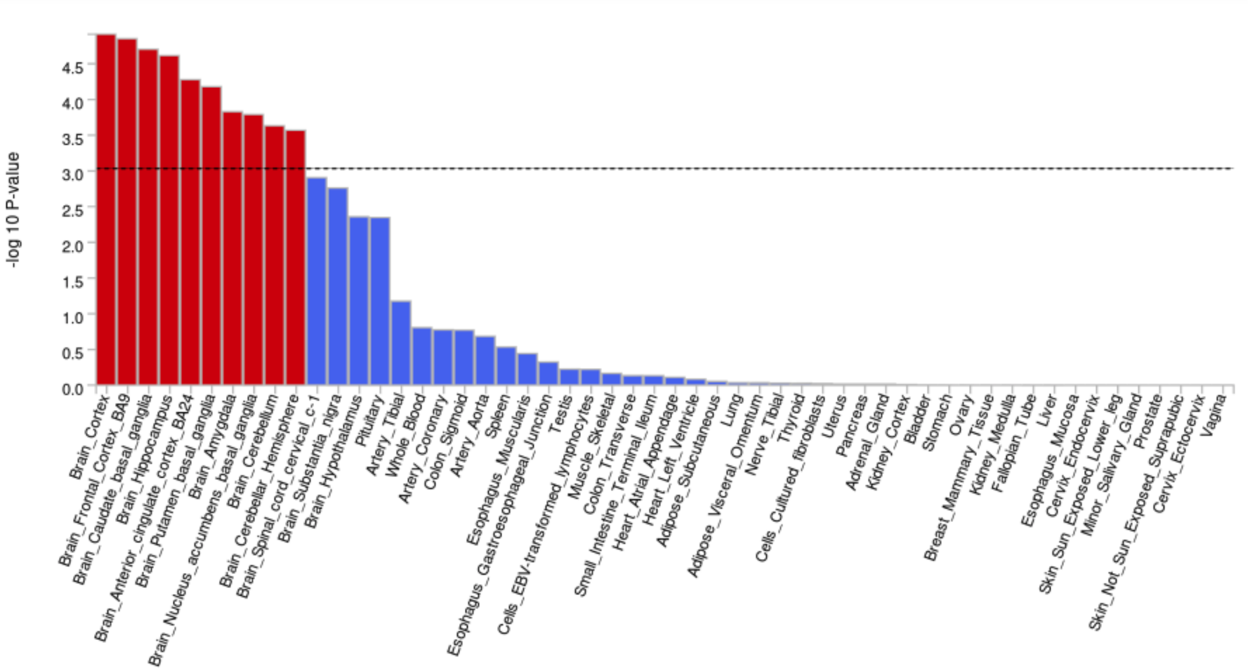


**Figure 10:** Top 100 Genes for PSYshared from gene-based analyses

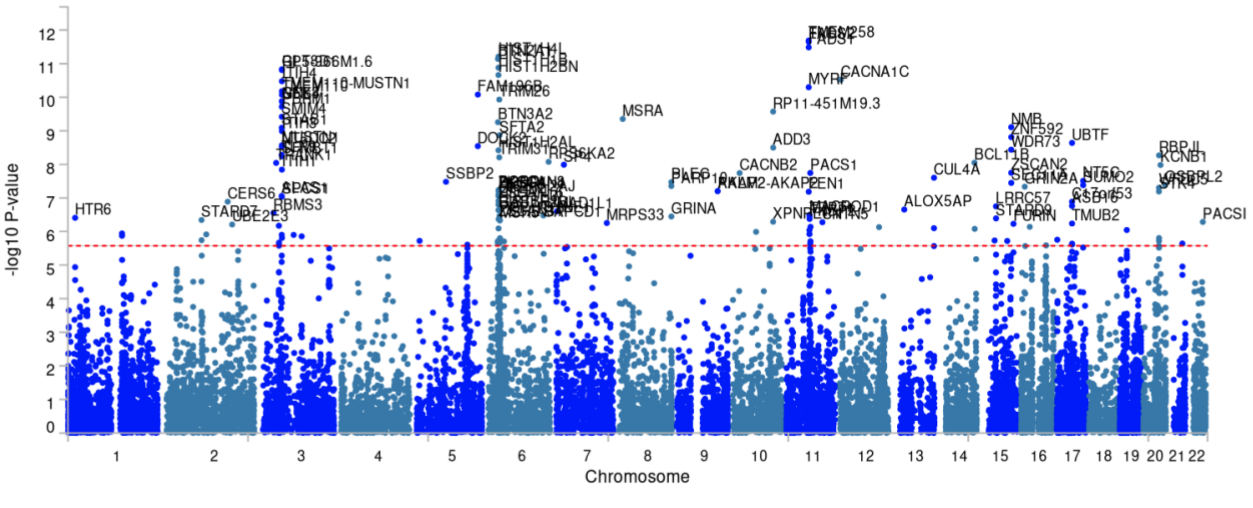

**Figure 11:** MAGMA Tissue Expression Analysis - PSYshared (GTEx v8 30 general tissue types)

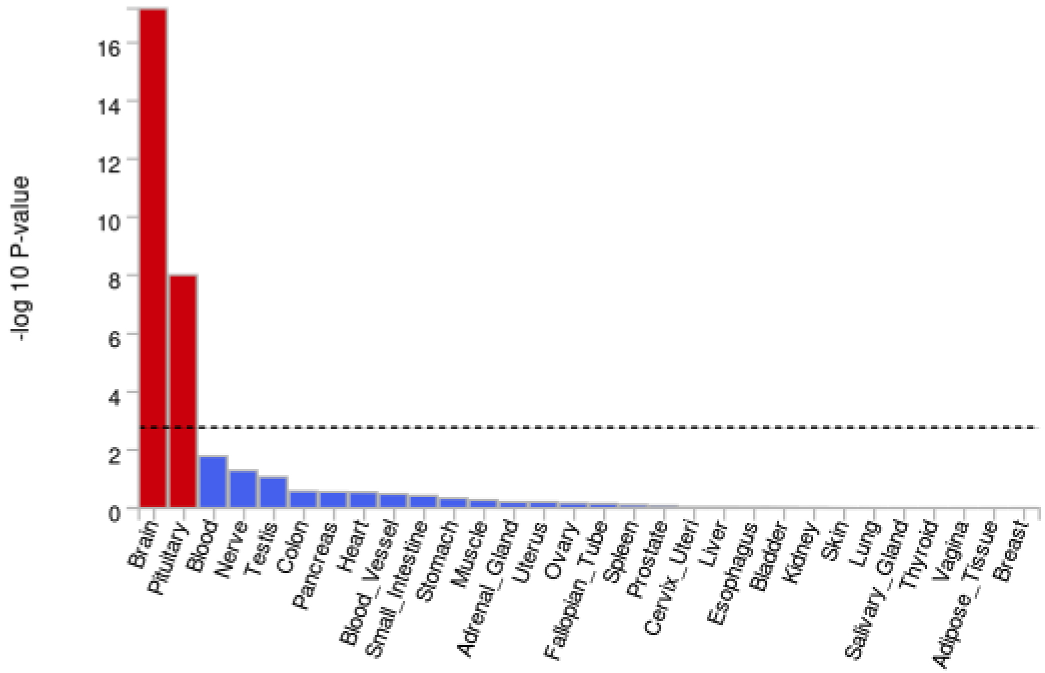


**Figure 12:** MAGMA Tissue Expression Analysis - PSYshared (GTEx v8 53 tissue types)

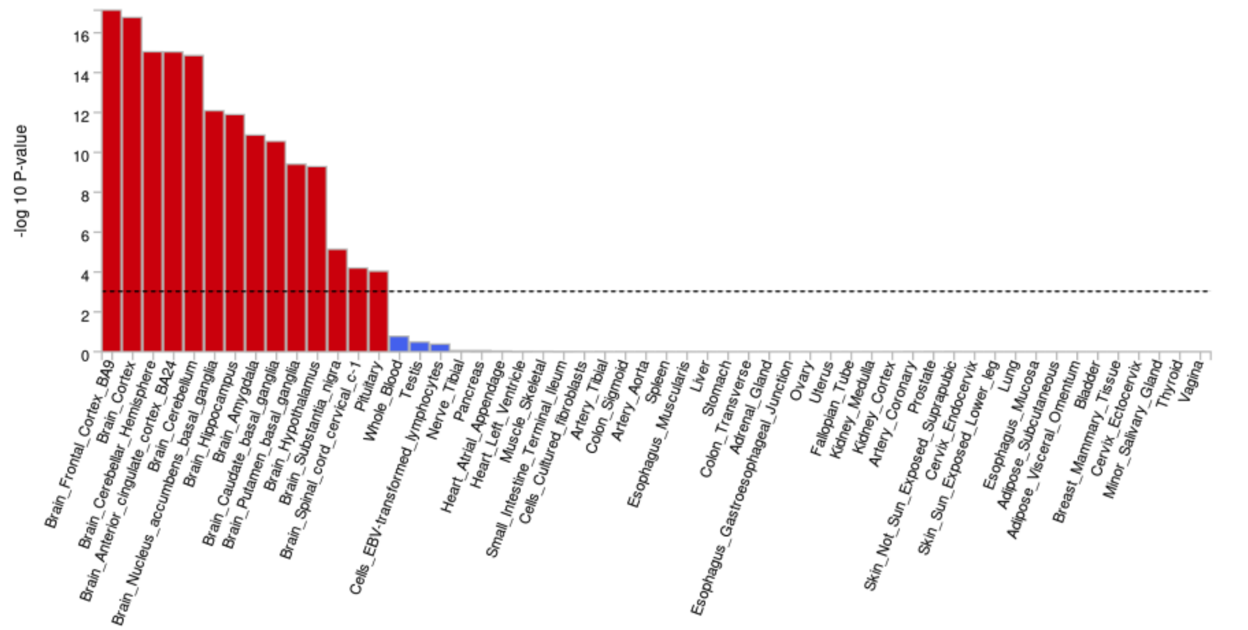

**Figure 13:** MR scatter plot schizophrenia on EA

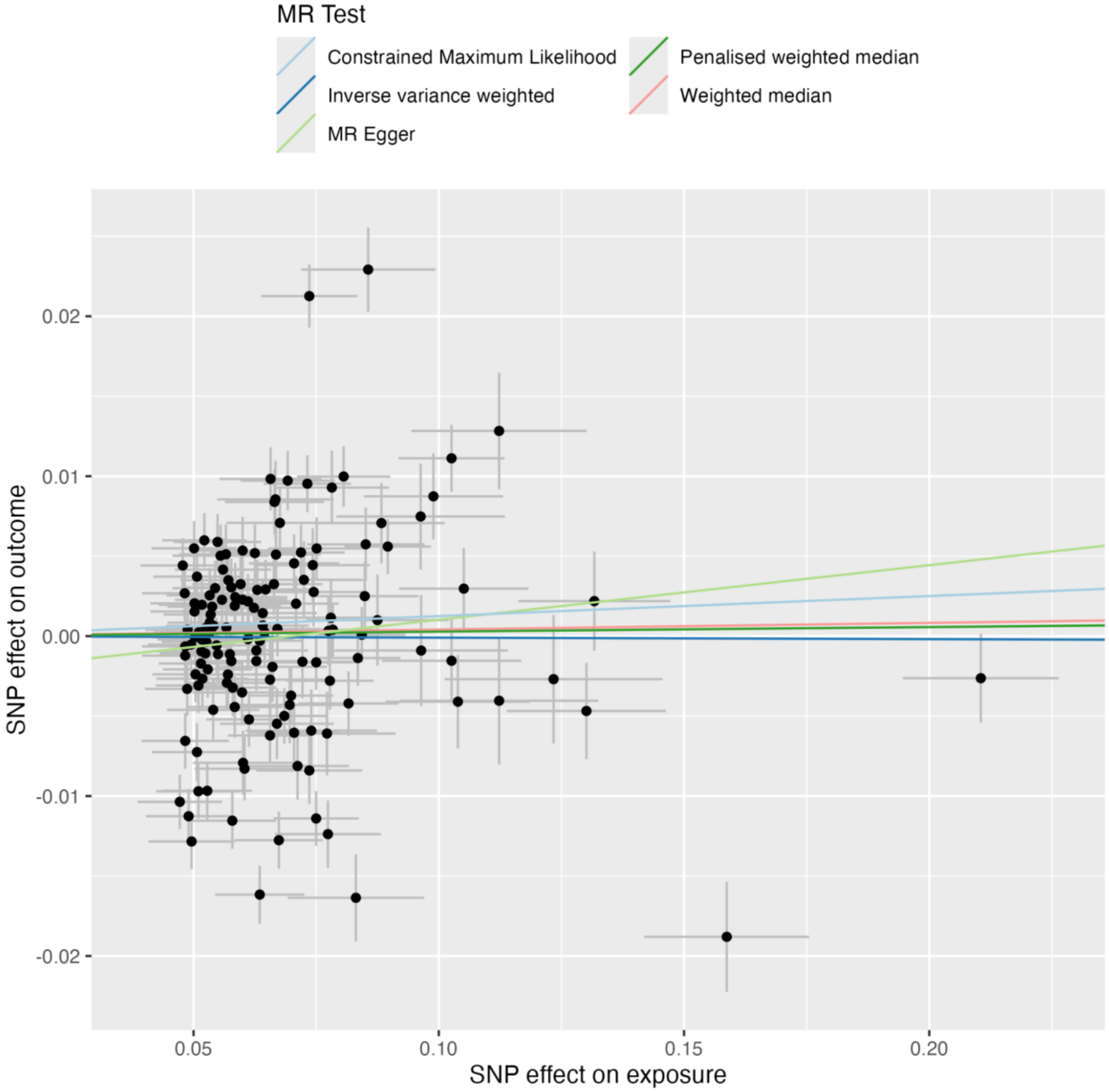

**Figure 14:** MR leave-one-out plot schizophrenia on EA

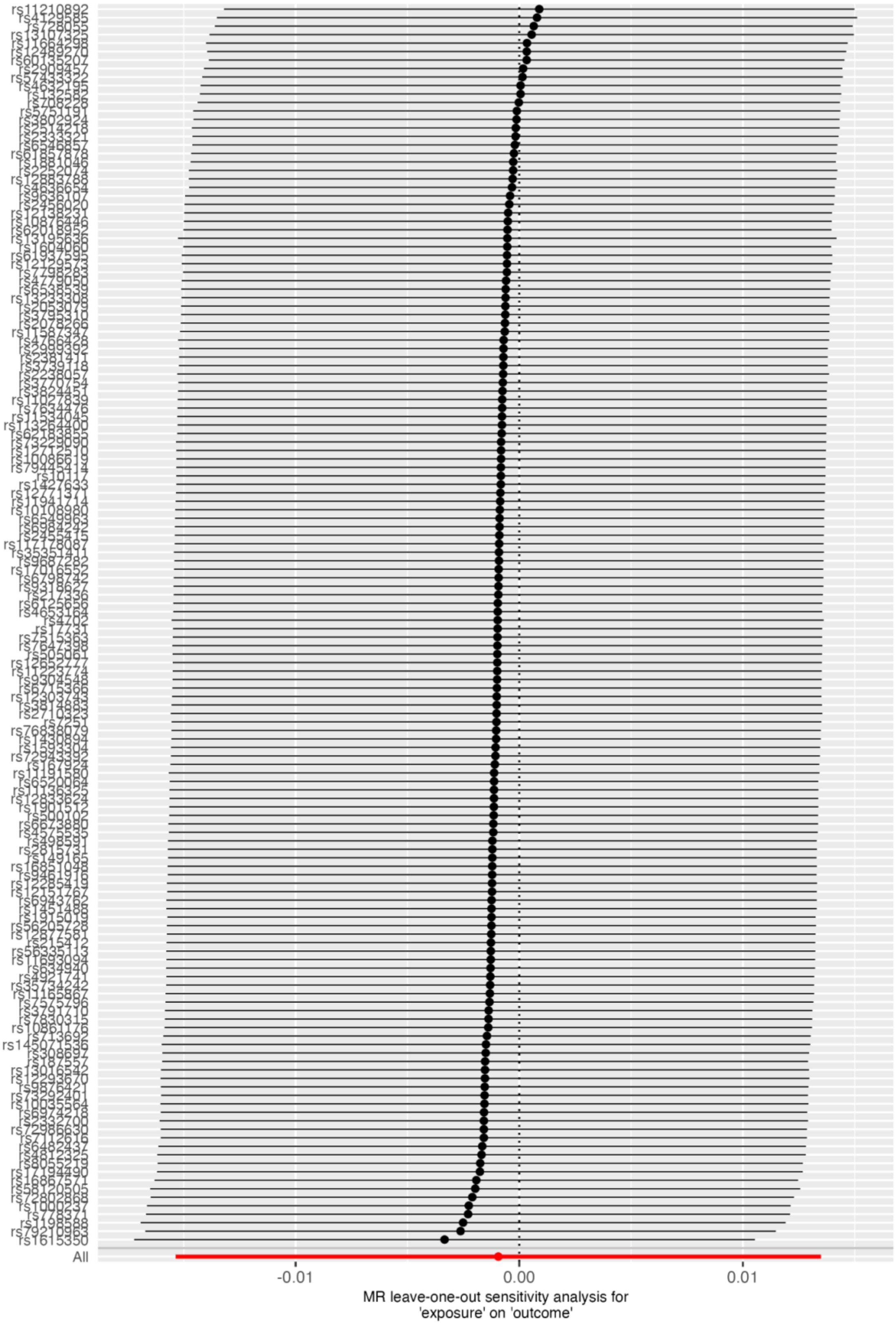

**Figure 15:** MR scatter plot Bipolar on EA

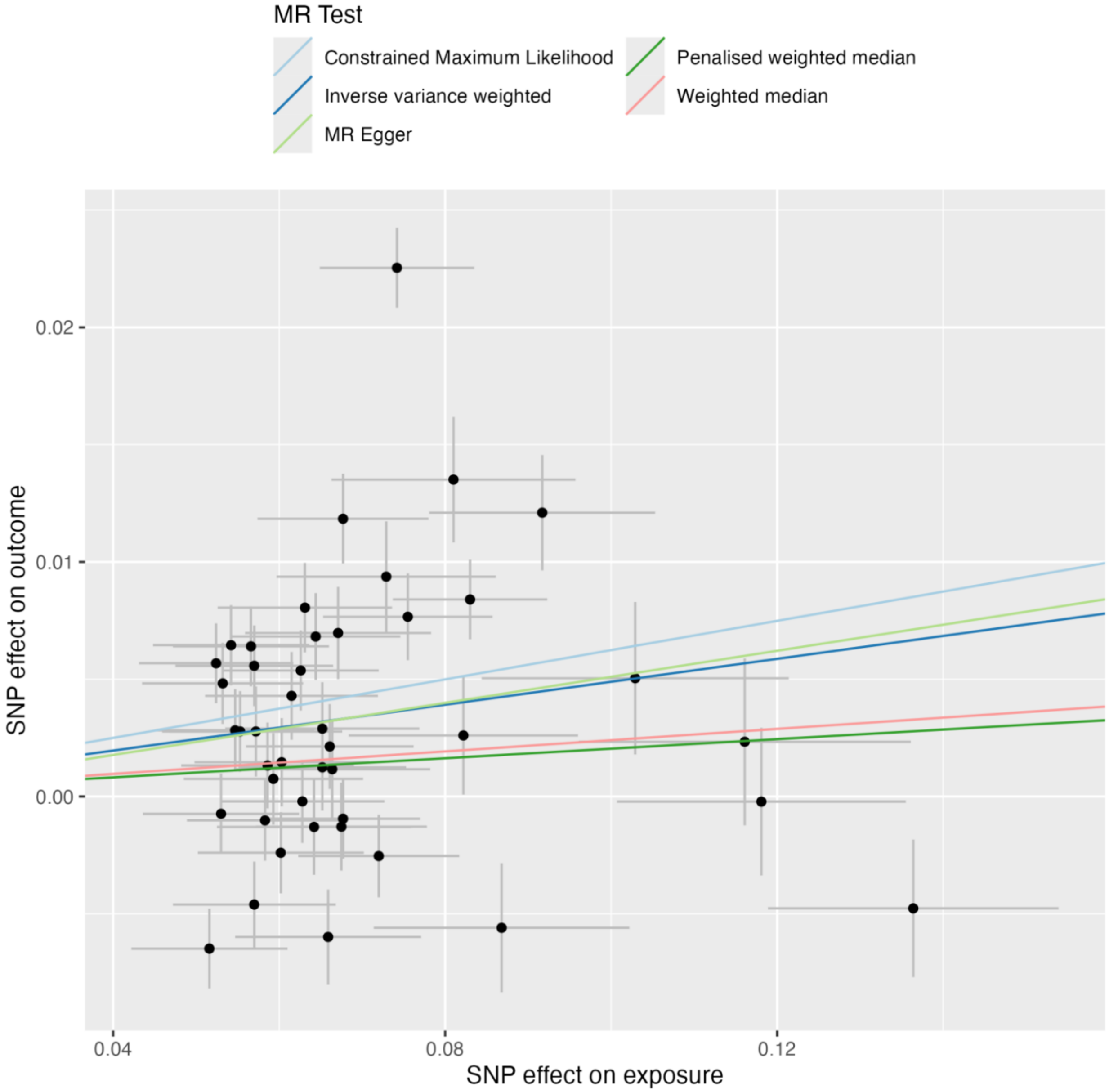

**Figure 16:** MR leave-one-out plot Bipolar on EA

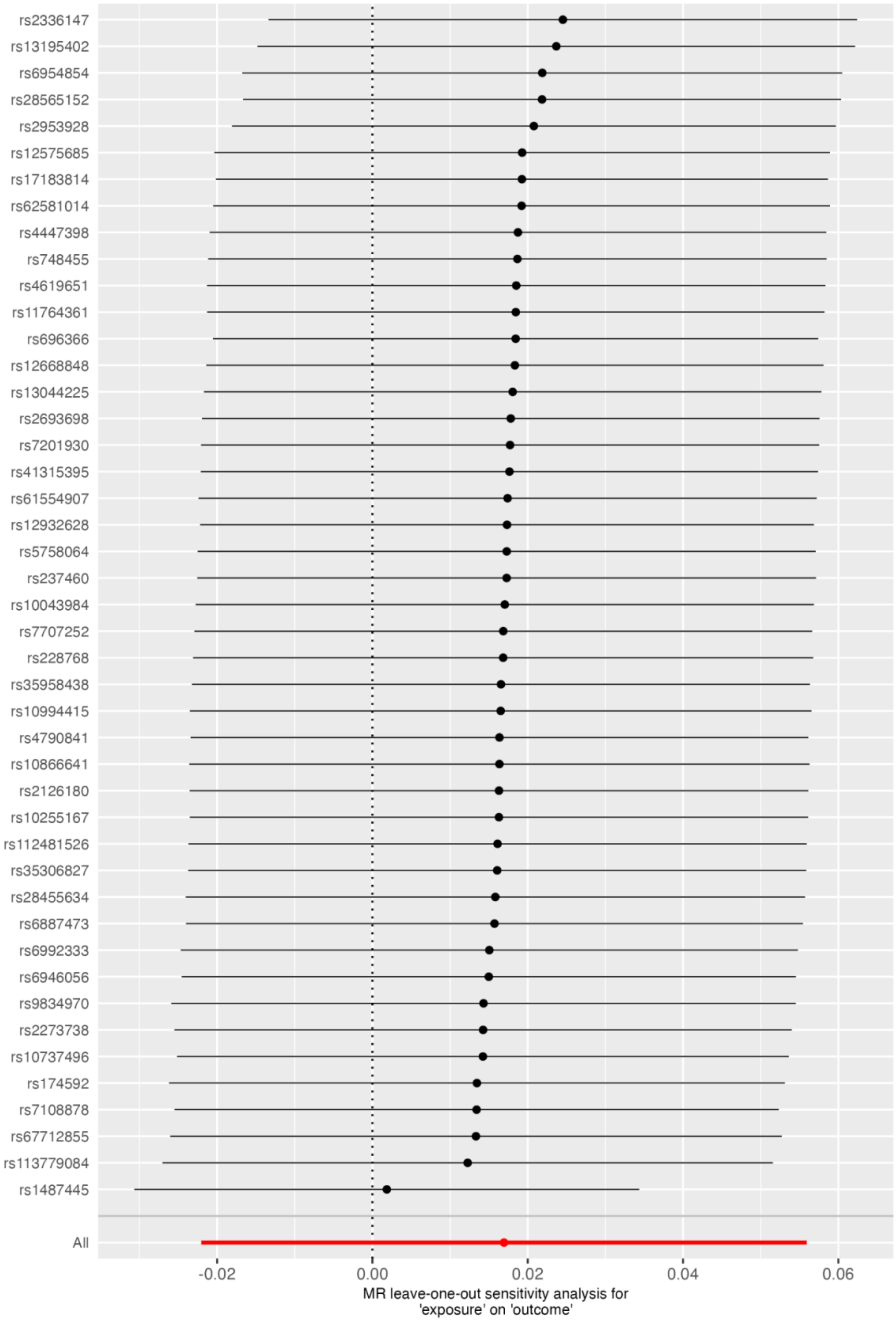

**Figure 17:** MR scatter plot SZspecific on EA

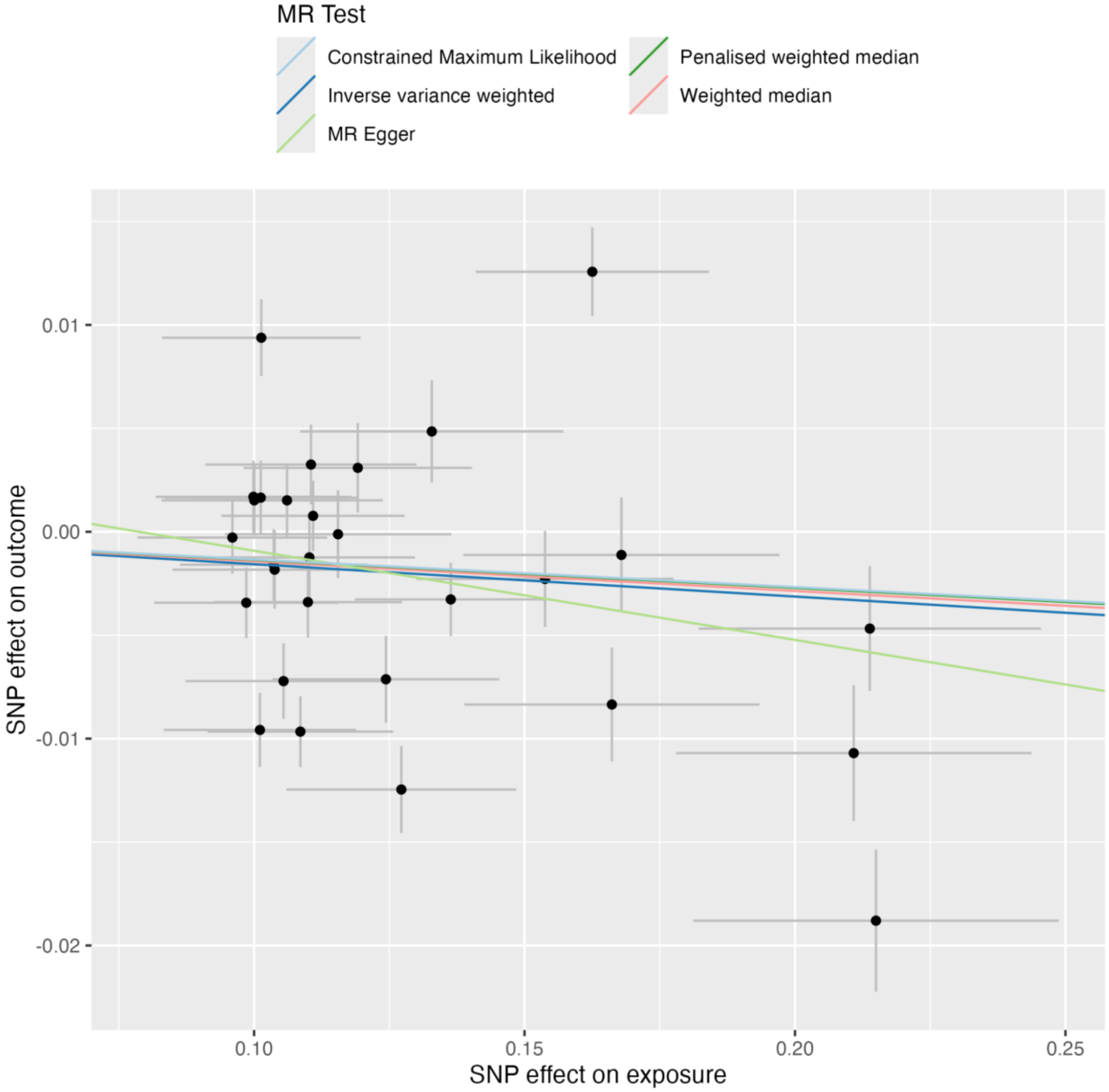

**Figure 18:** MR leave-one-out plot SZspecific on EA

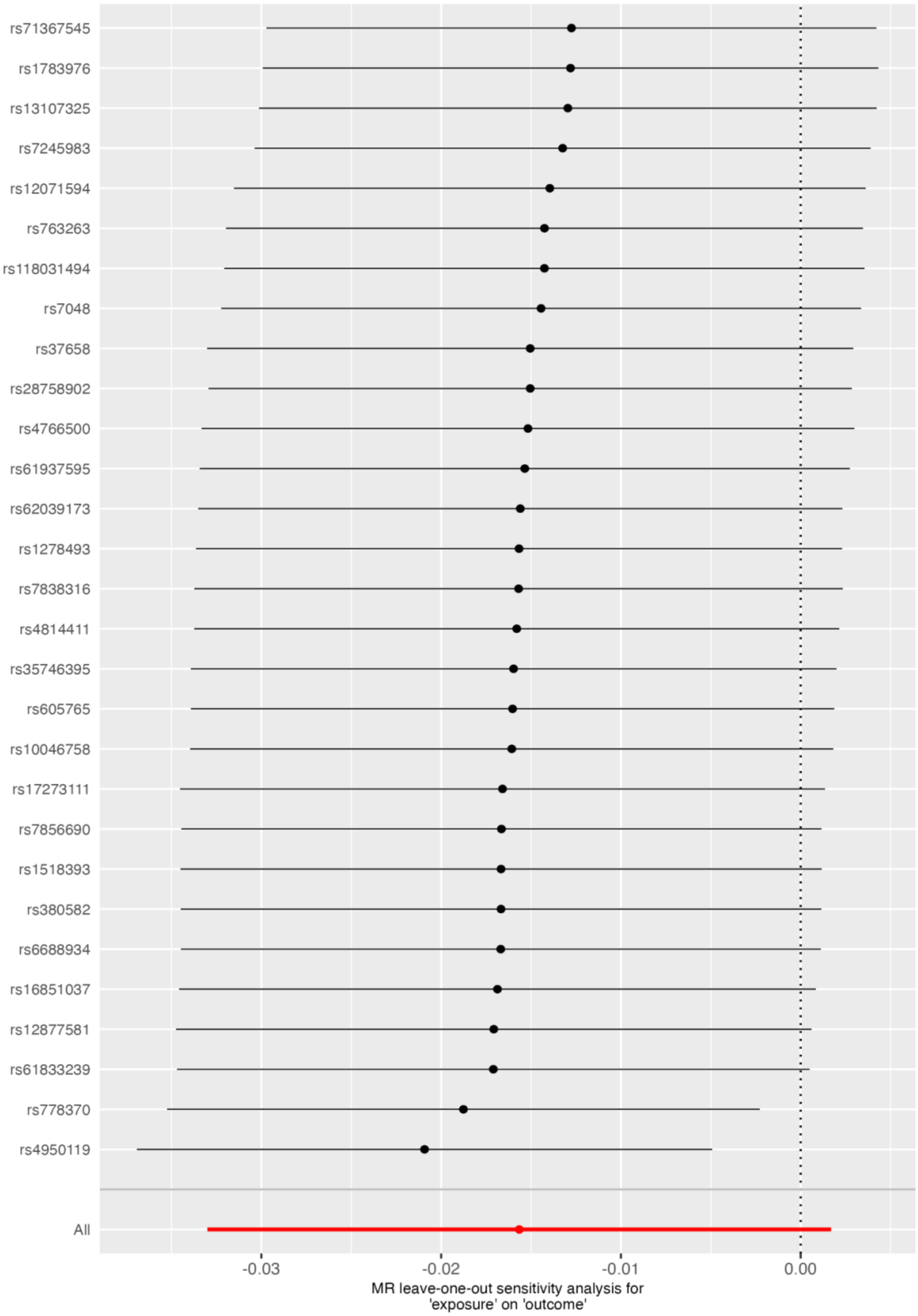

**Figure 19:** MR scatter plot PSYshared on EA

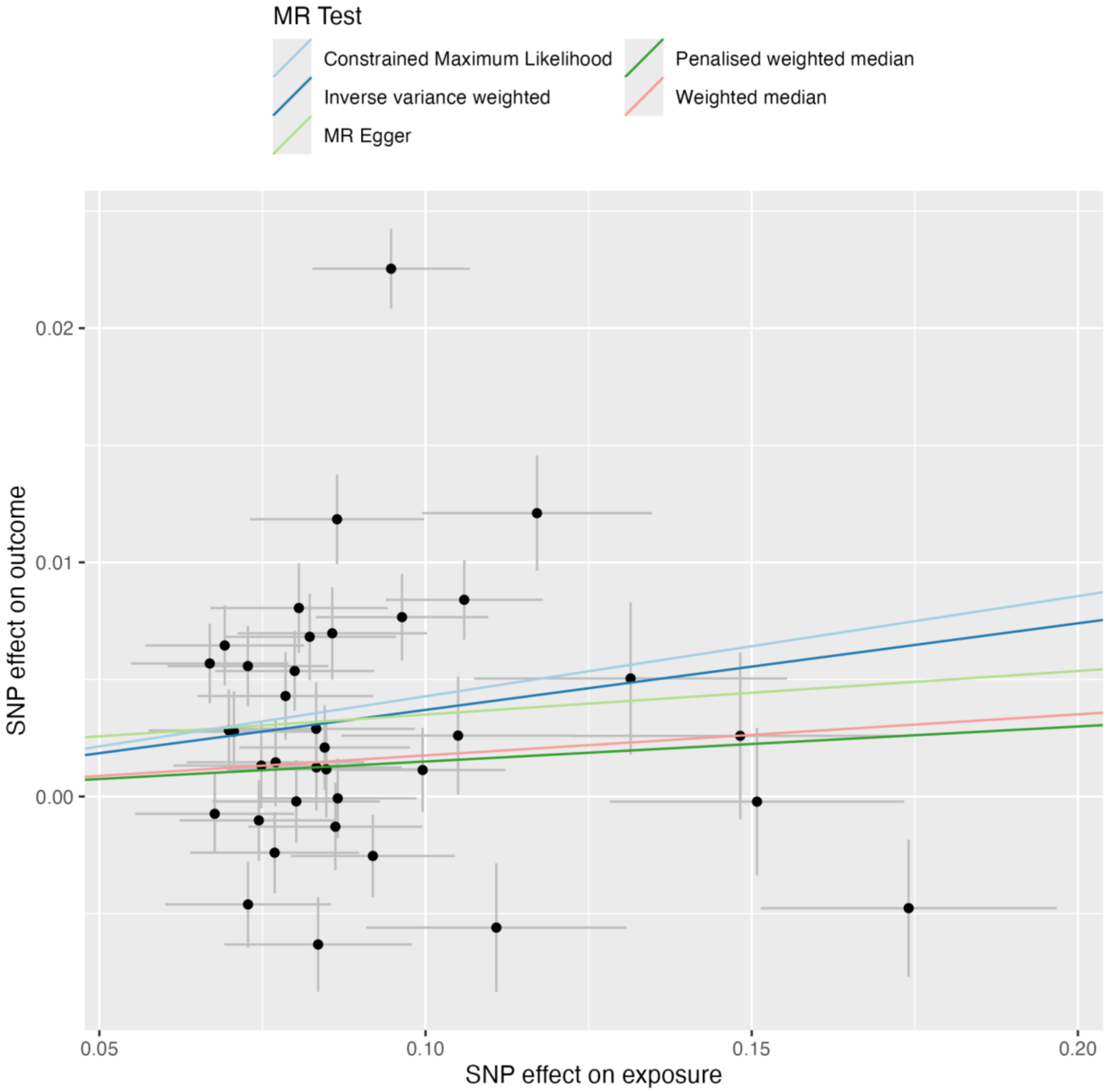

**Figure 20:** MR leave-one-out plot PSYshared on EA

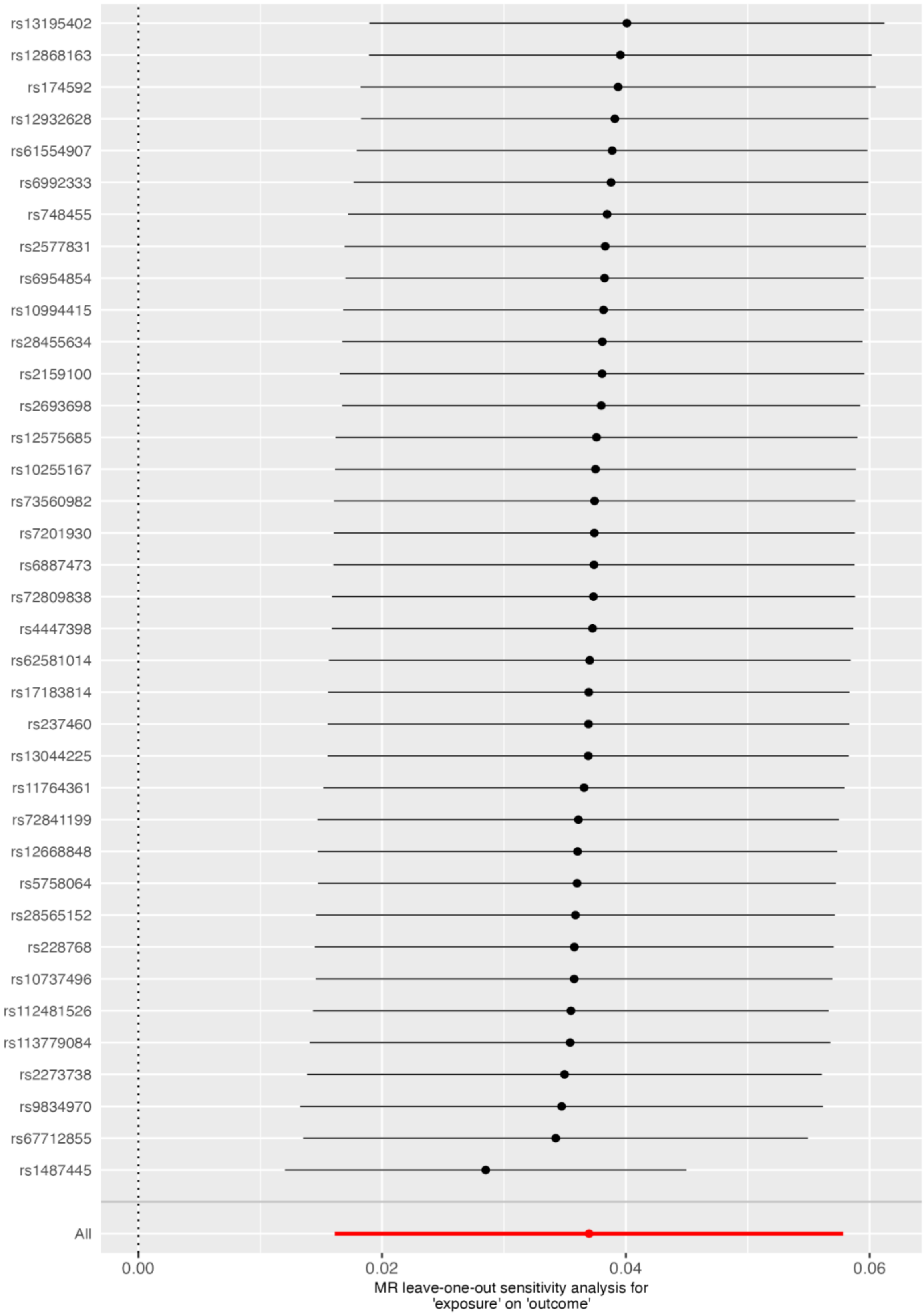

**Figure 21:** MR scatter plot Schizophrenia on IQ

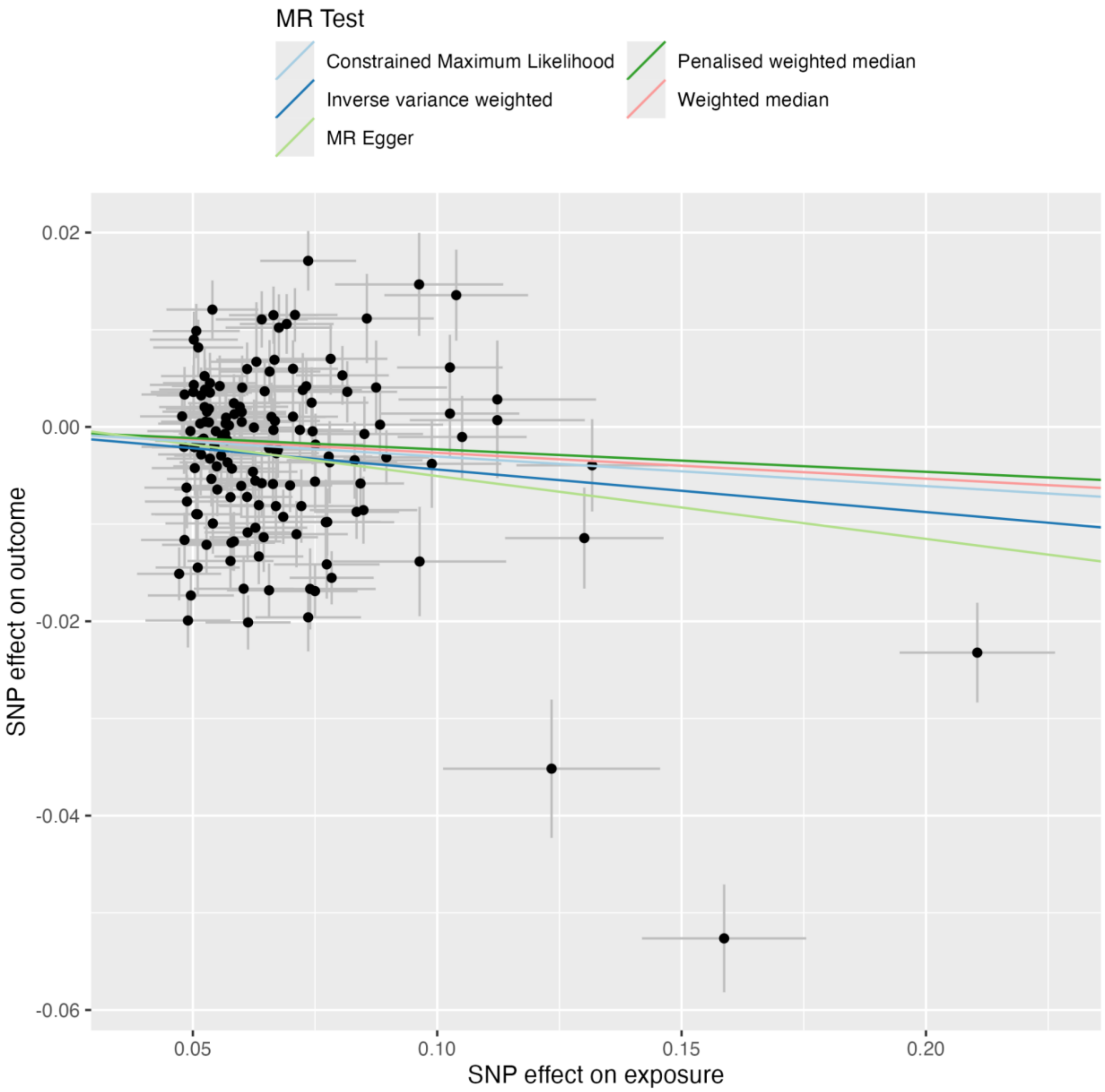

**Figure 22:** MR leave-one-out plot Schizophrenia on IQ

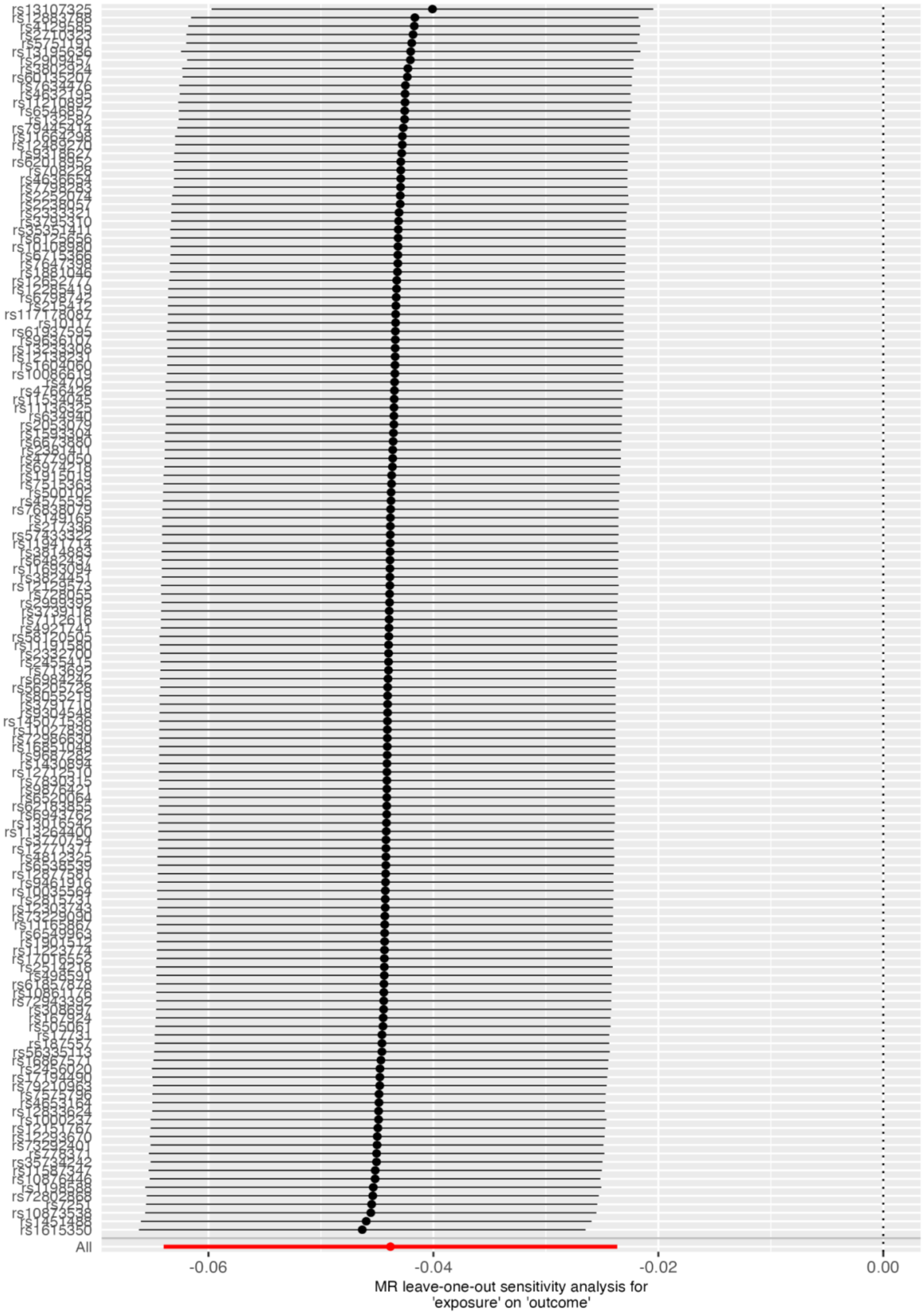

**Figure 23:** MR scatter plot Bipolar on IQ

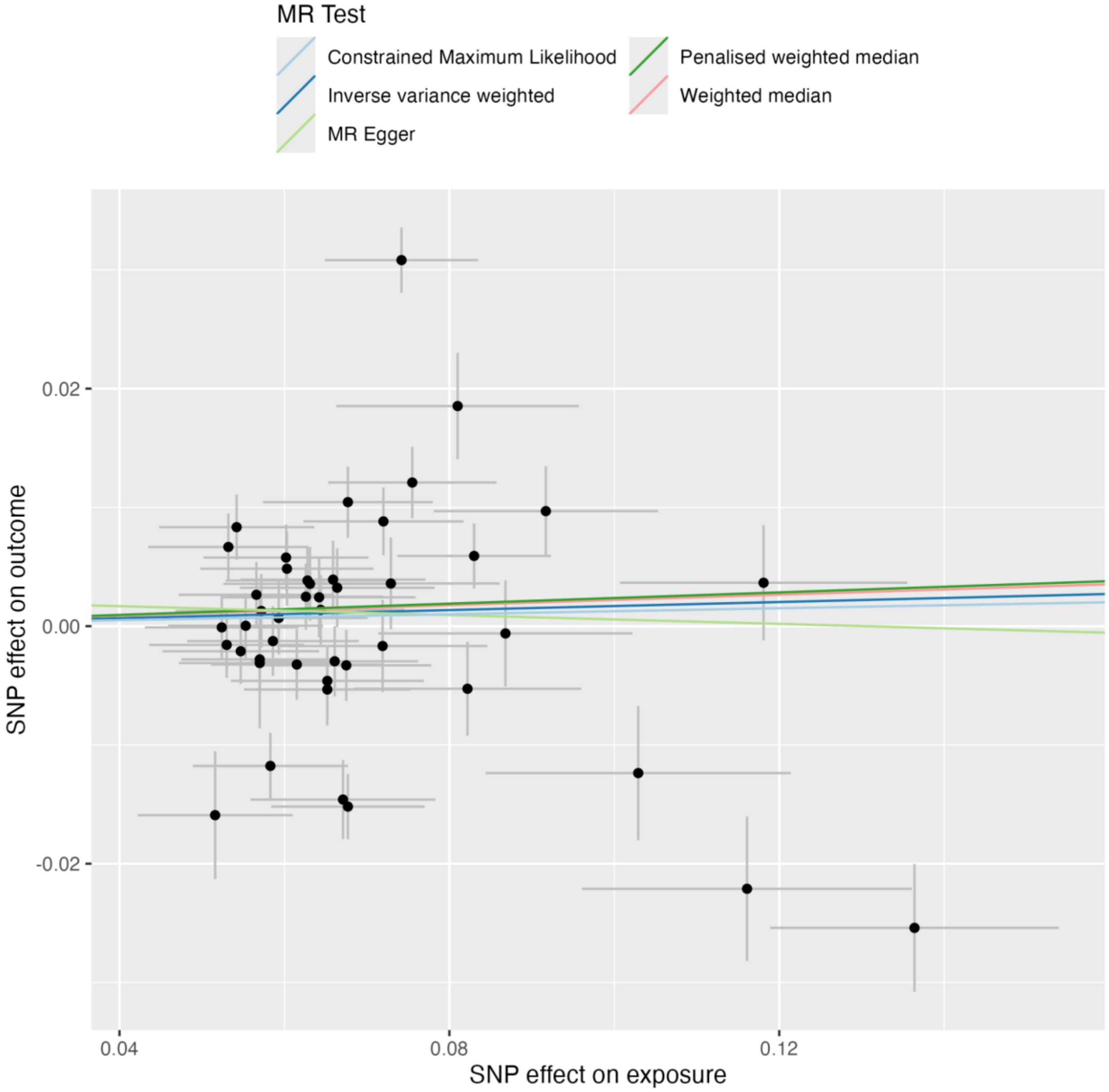

**Figure 24:** MR leave-one-out plot Bipolar on IQ

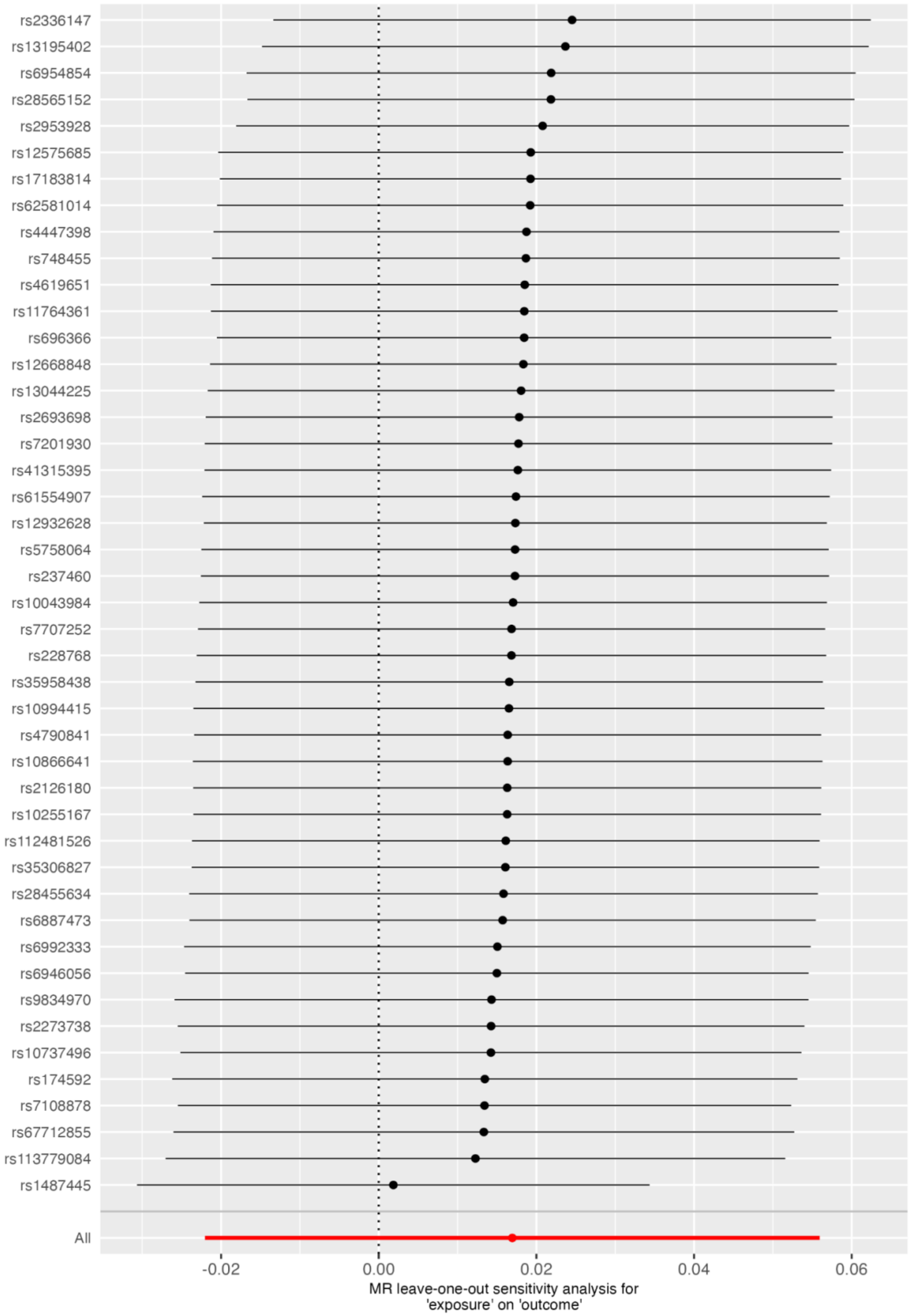

**Figure 25:** MR scatter plot SZspecific on IQ

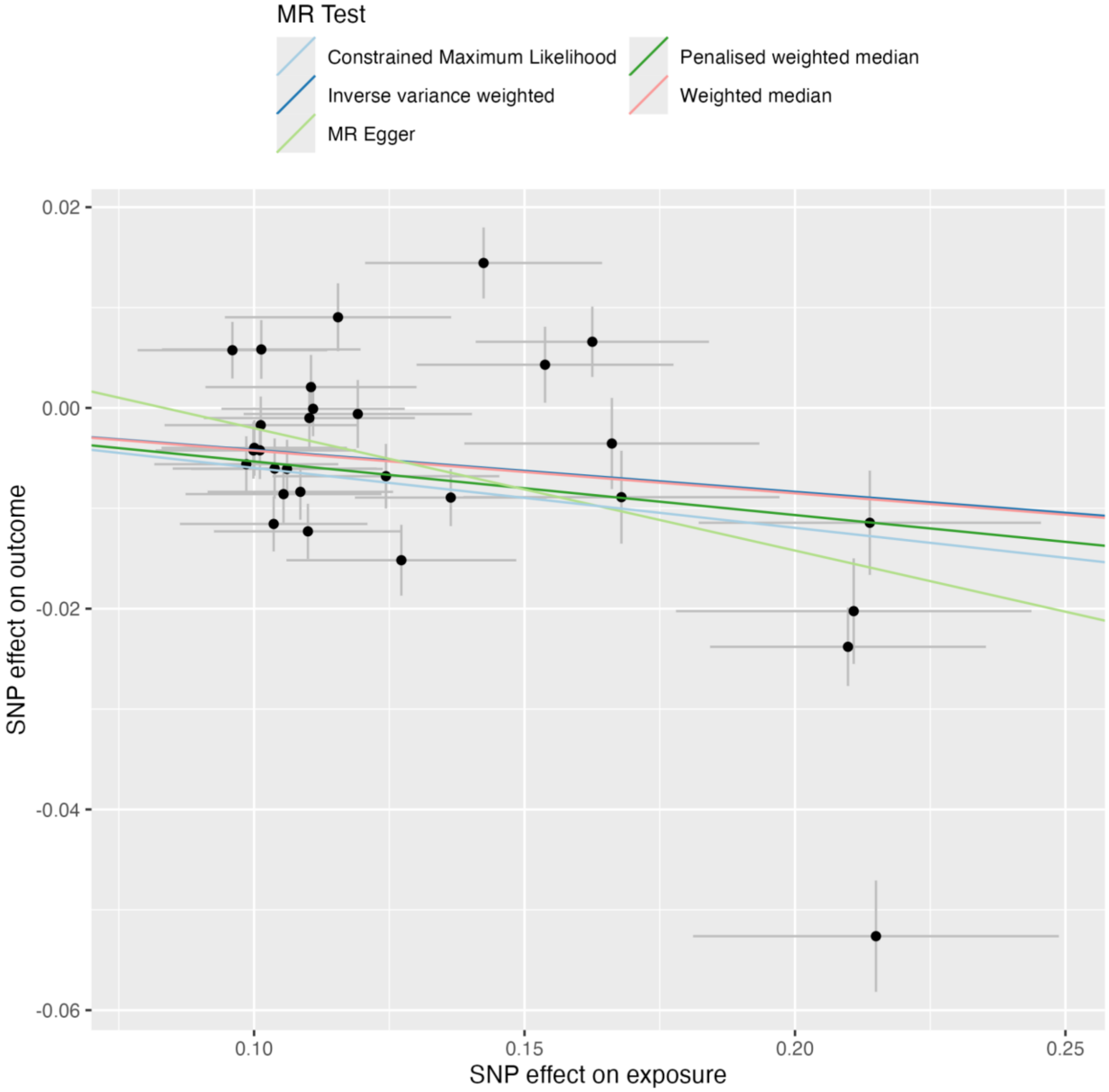

**Figure 26:** MR leave-one-out plot SZspecific on IQ

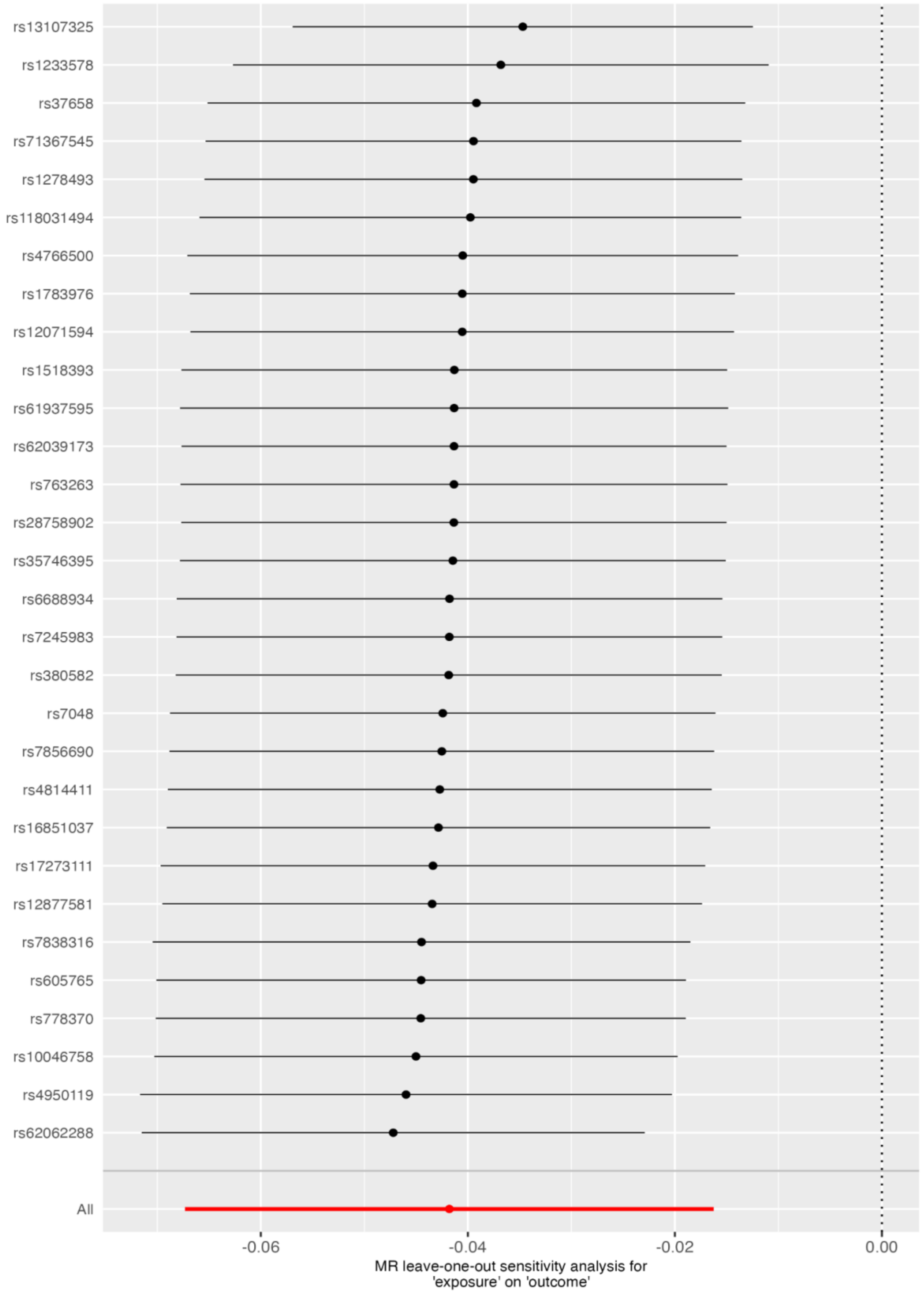

**Figure 27:** MR scatter plot PSYshared on IQ

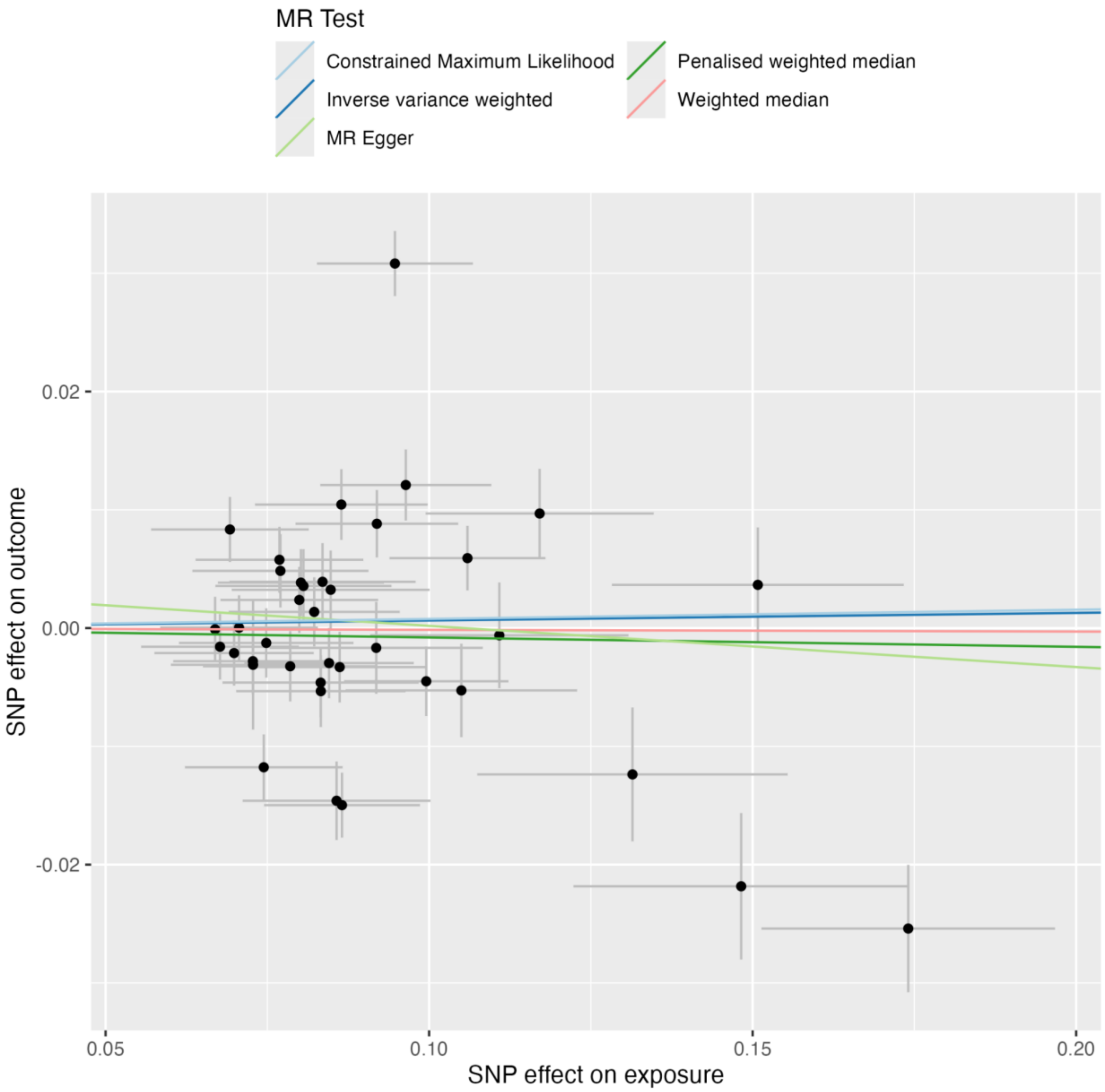

**Figure 28:** MR leave-one-out plot PSYshared on IQ

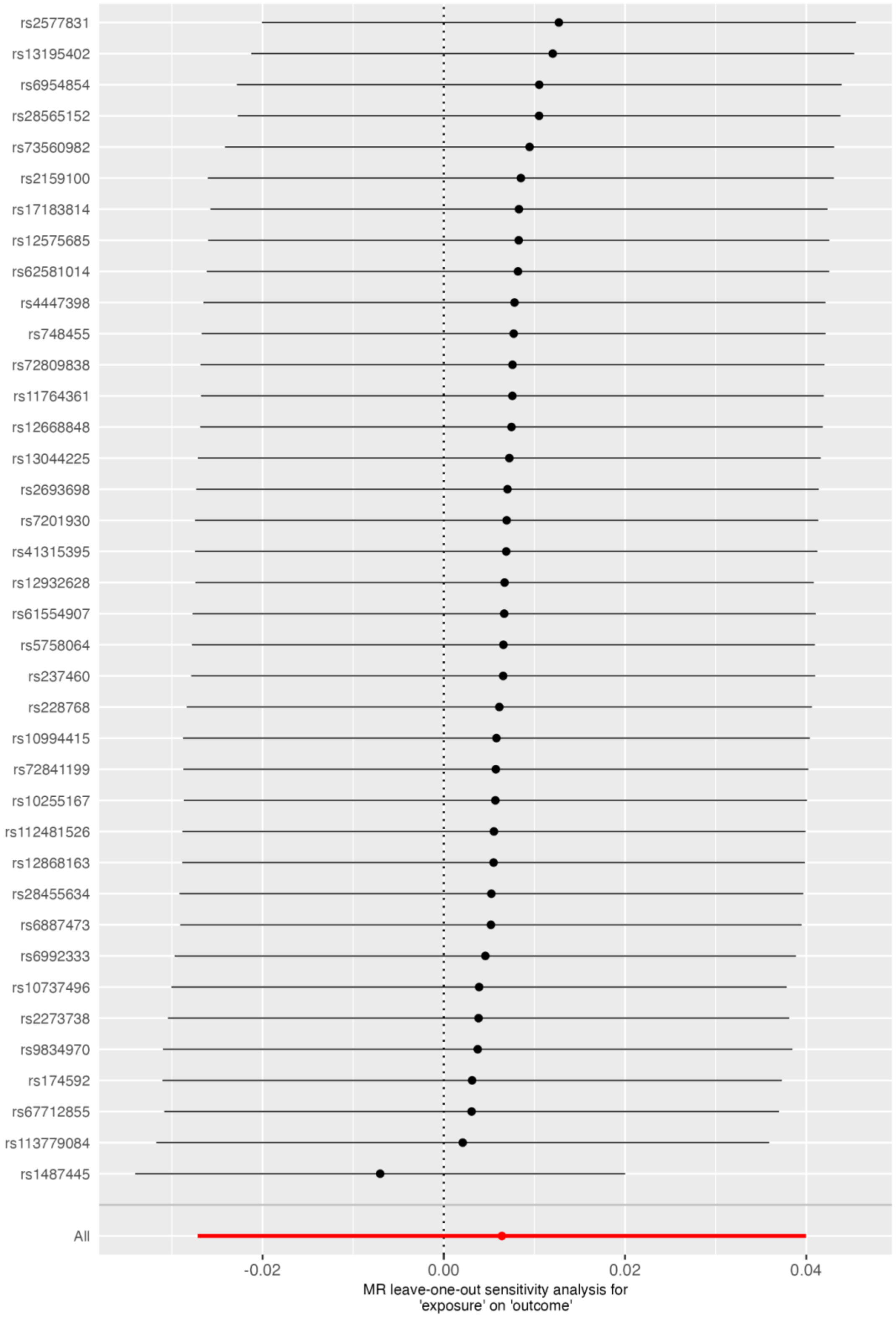

**Figure 29:** MR scatter plot EA on Schizophrenia

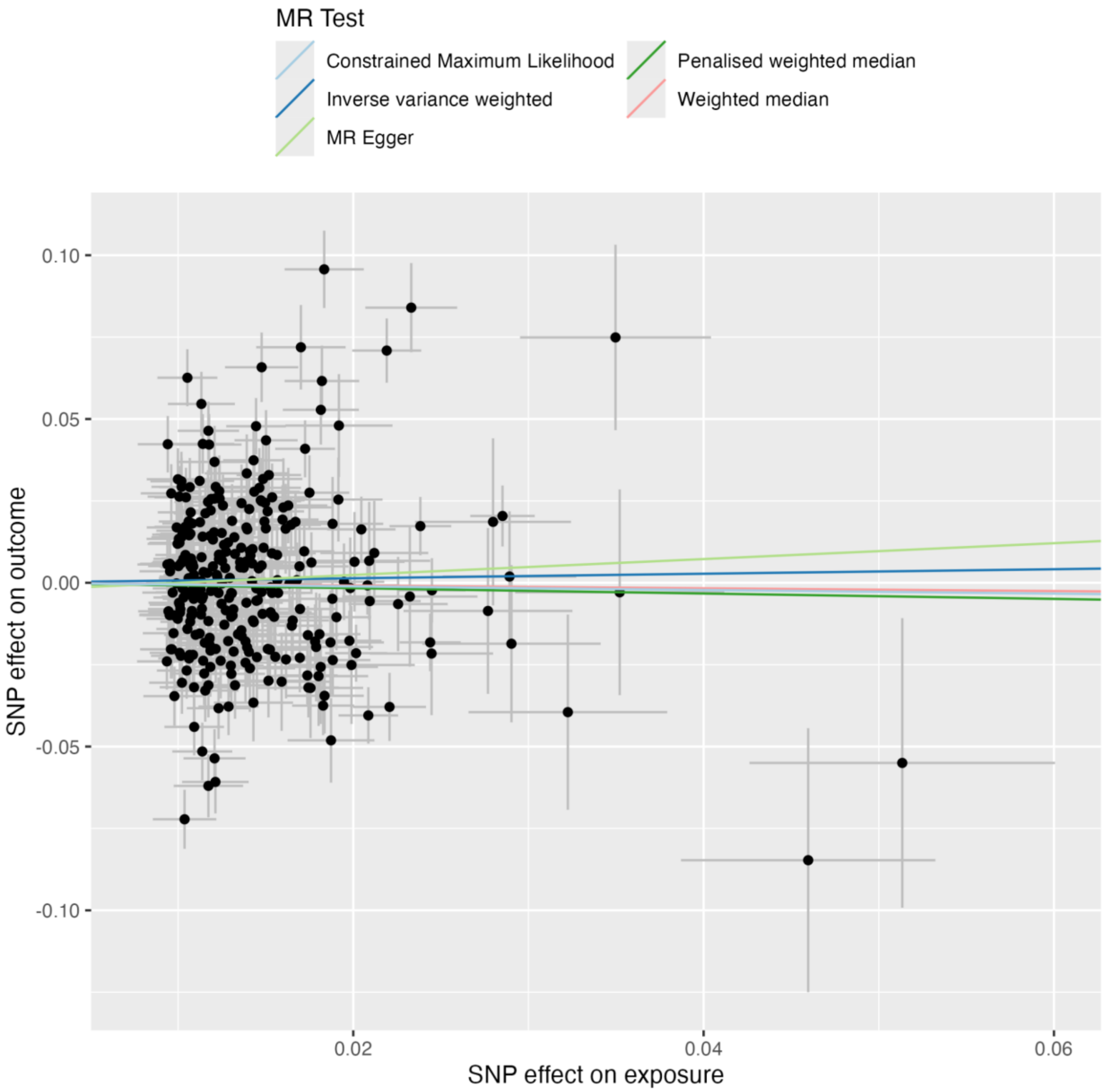

**Figure 30:** MR leave-one-out plot EA on Schizophrenia

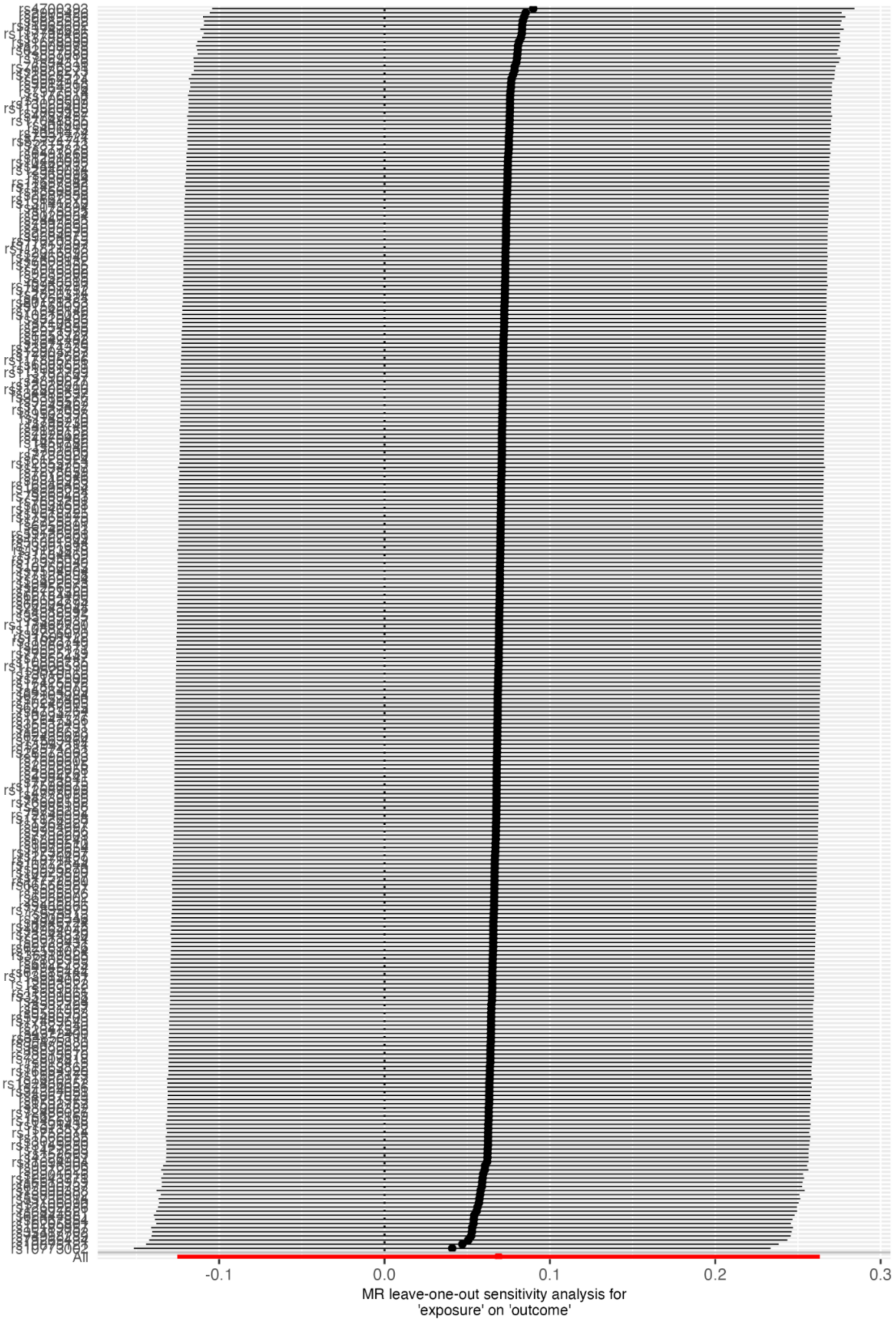

**Figure 31:** MR scatter plot EA on Bipolar

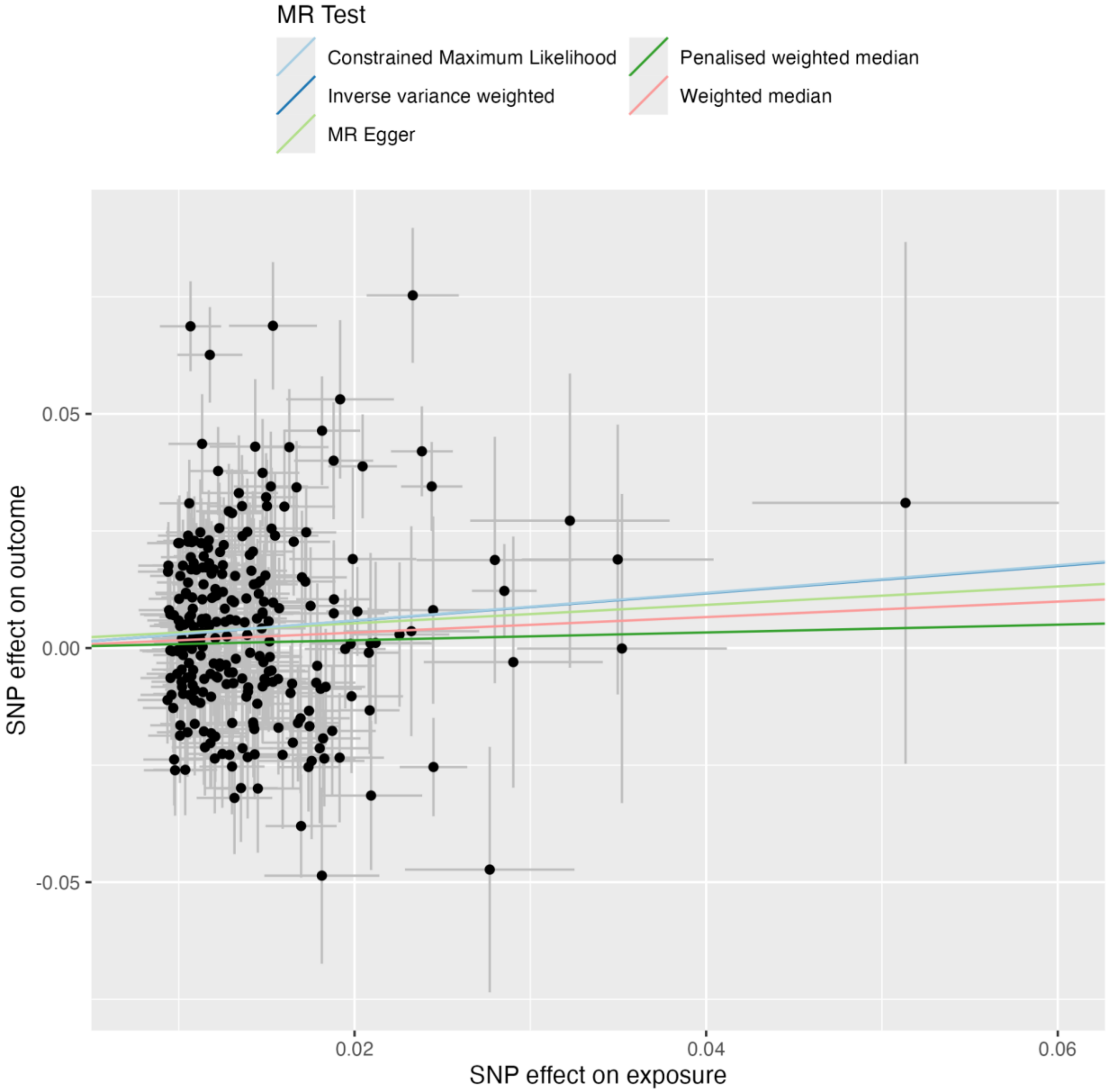

**Figure 32:** MR leave-one-out plot EA on Bipolar

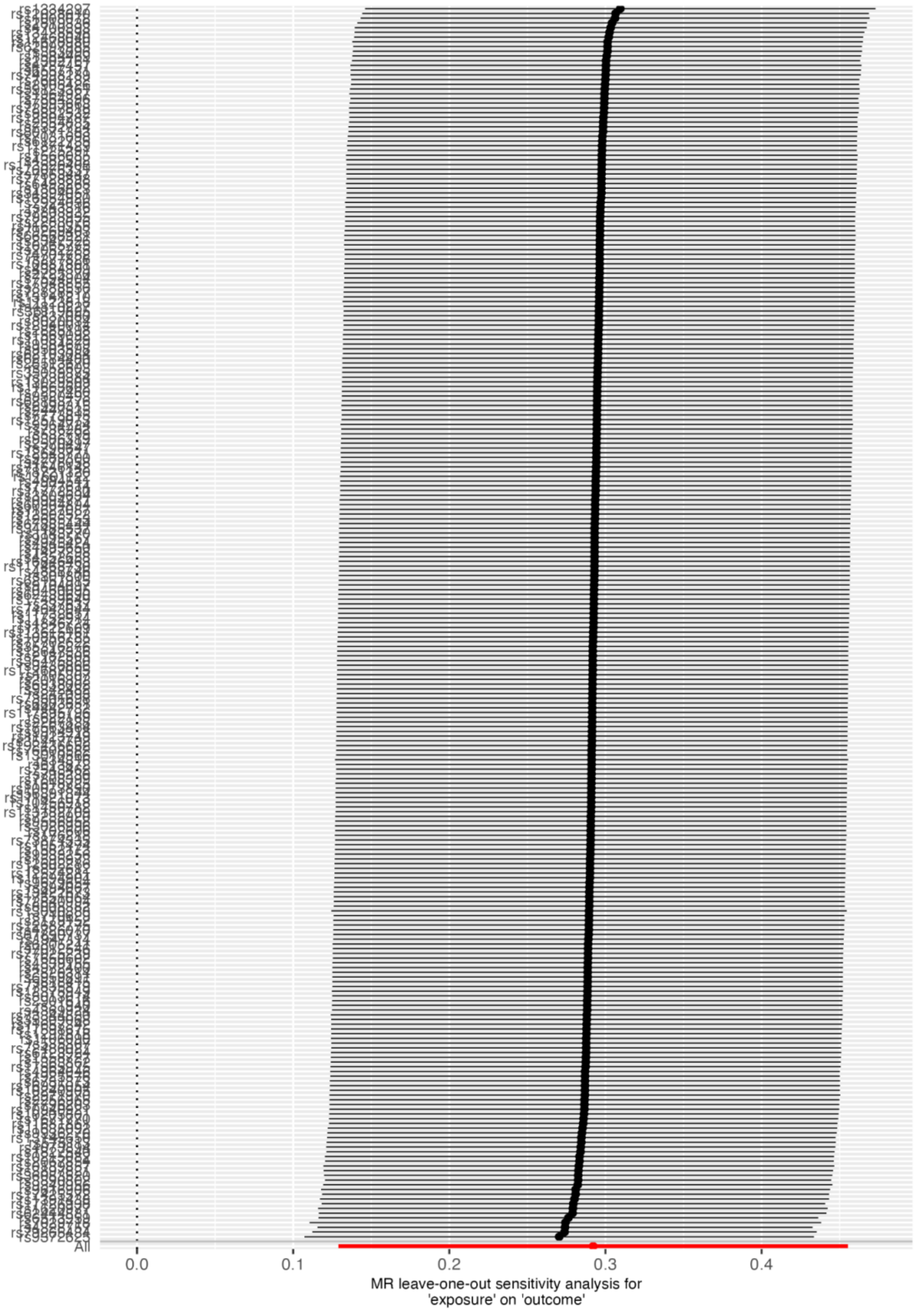

**Figure 33:** MR scatter plot EA on SZspecific

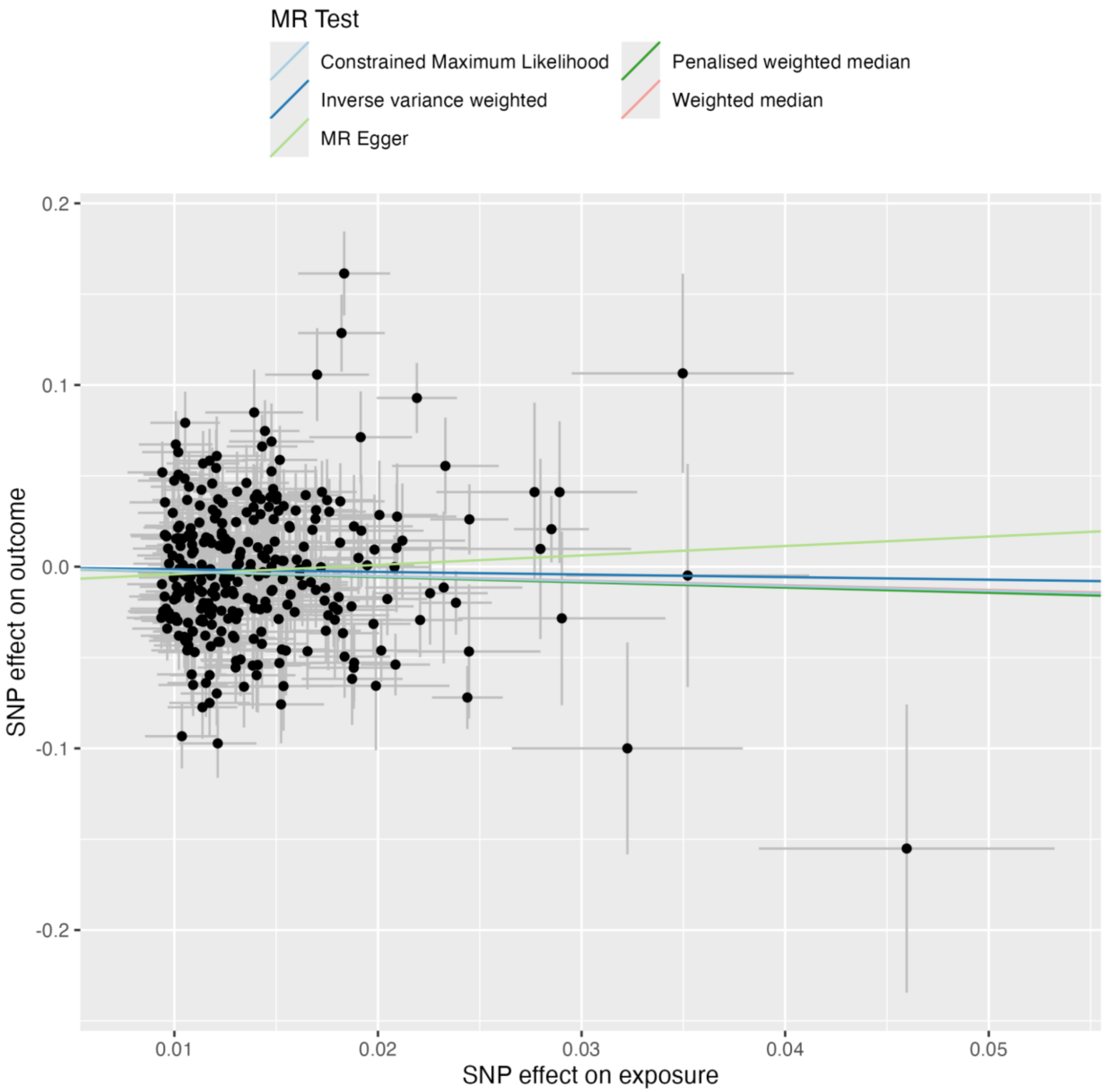

**Figure 34:** MR leave-one-out plot EA on SZspecific

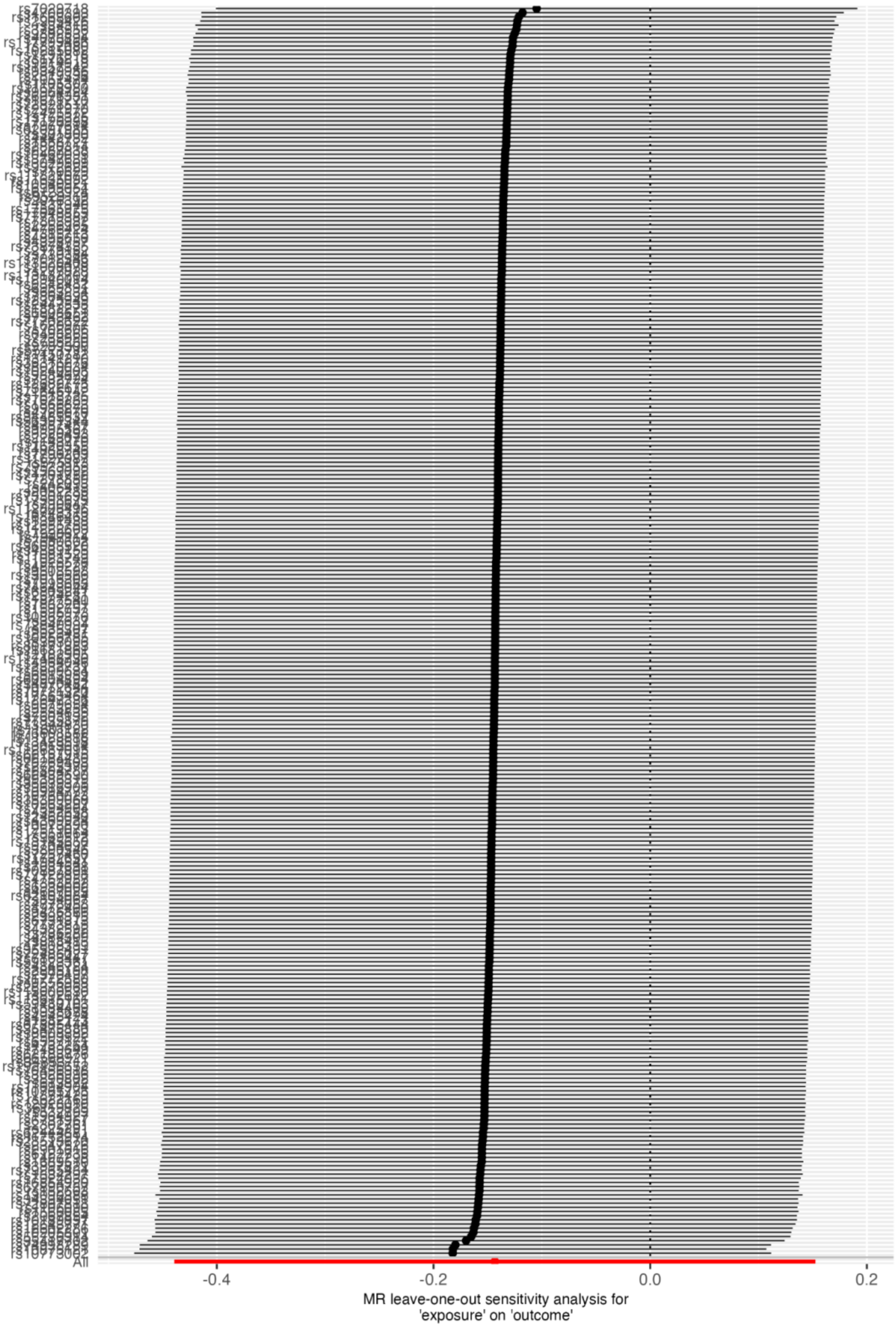

**Figure 35:** MR scatter plot EA on PSYshared

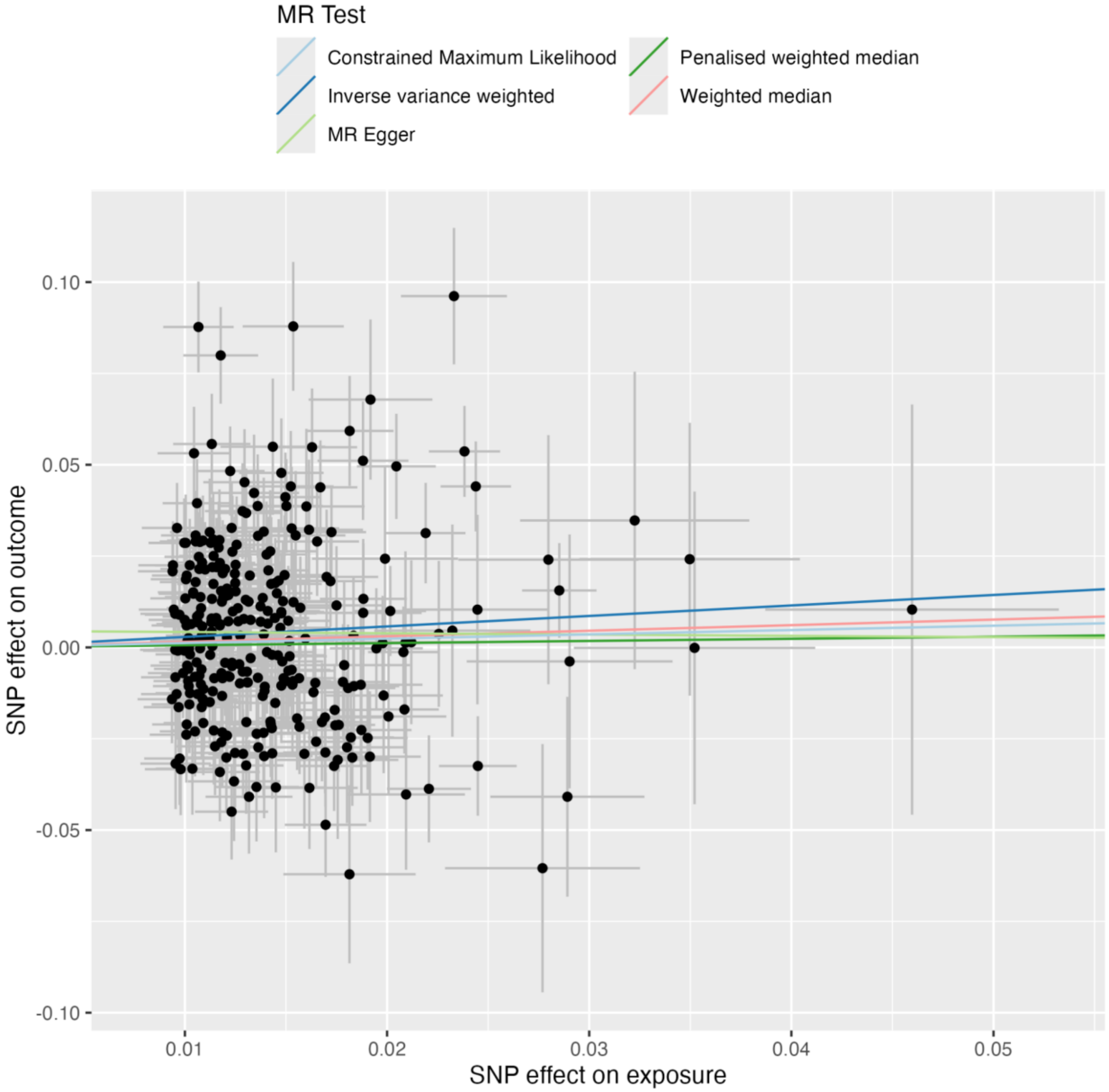

**Figure 36:** MR leave-one-out plot EA on PSYshared

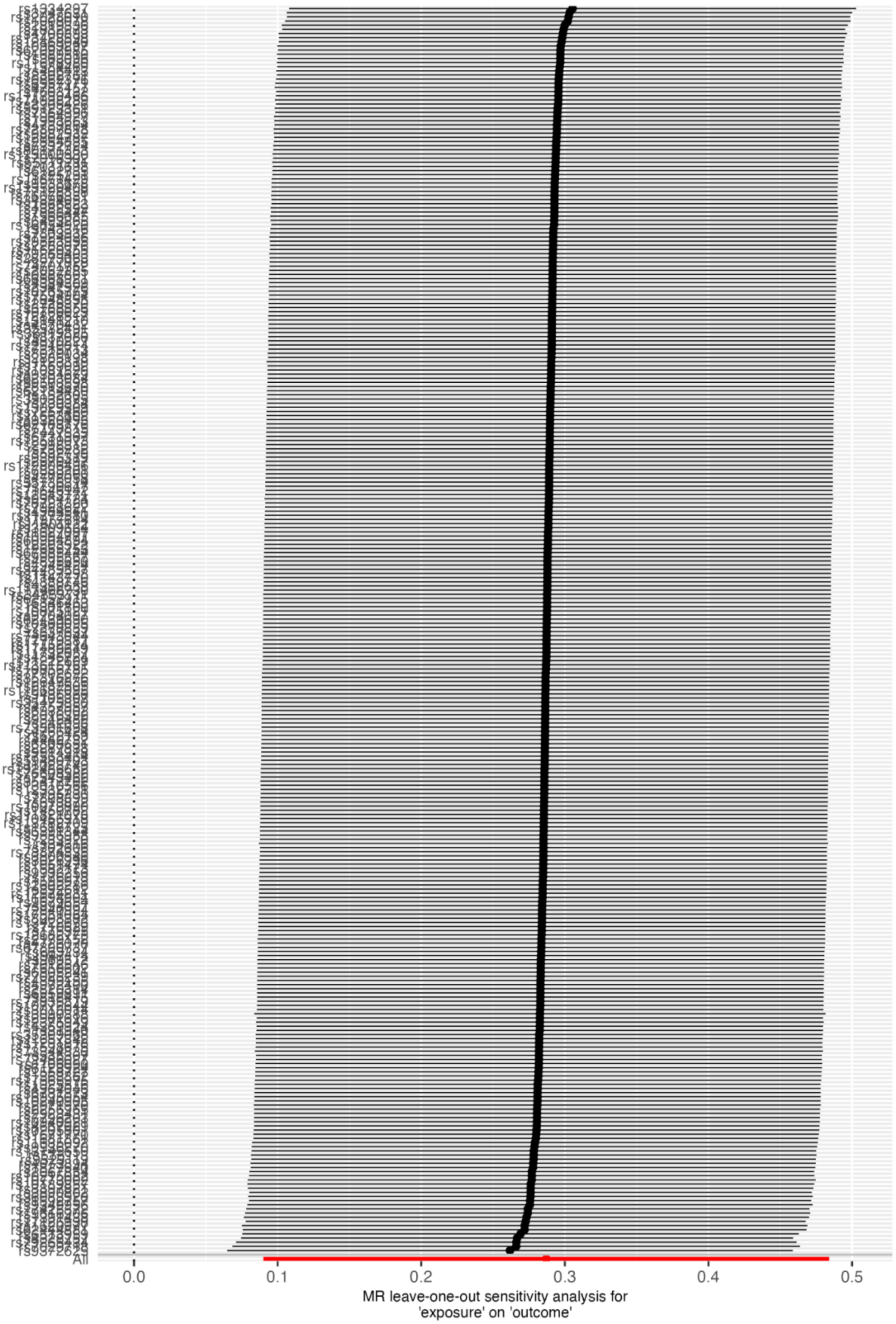

**Figure 37:** MR scatter plot IQ on Schizophrenia

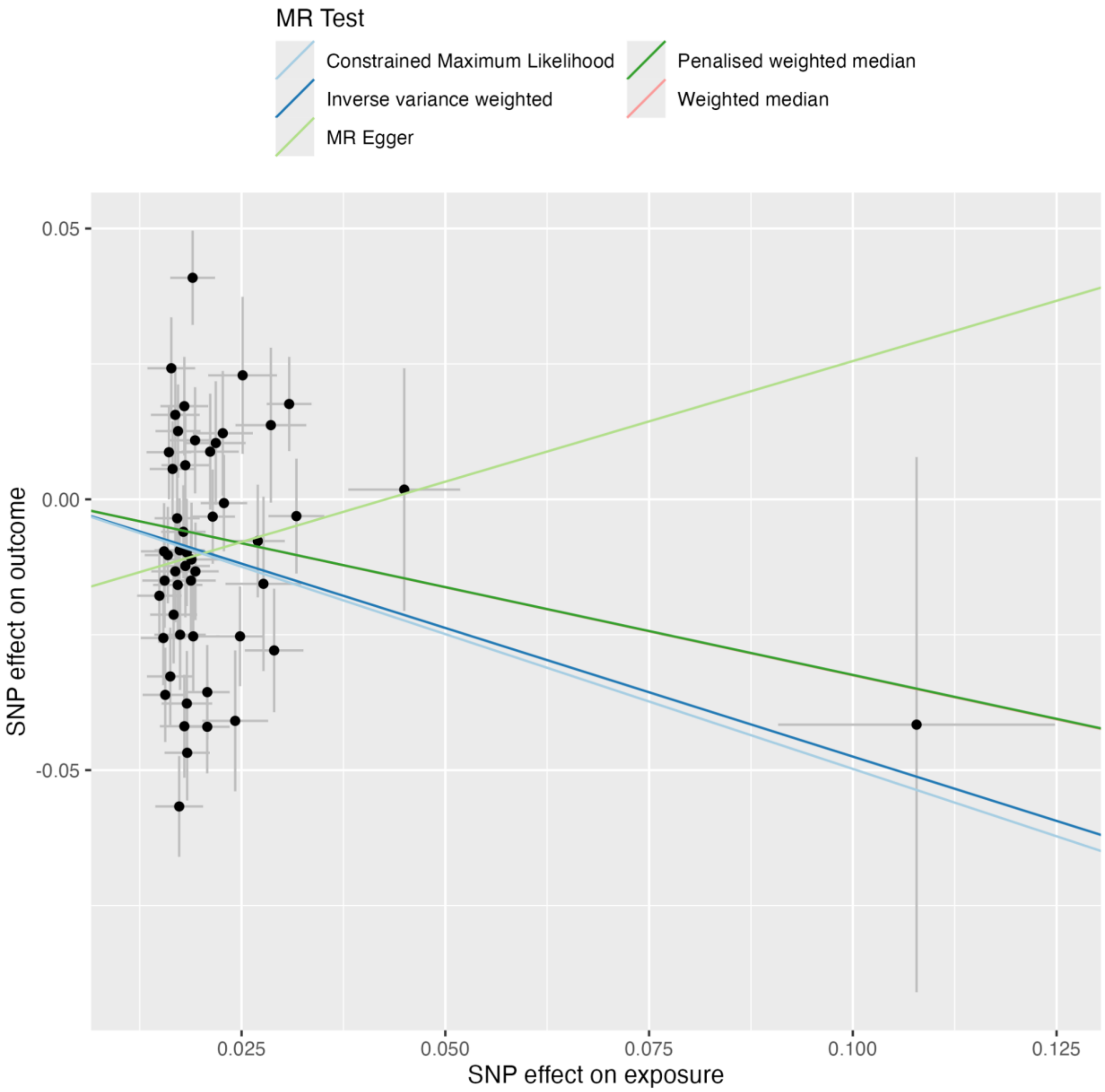

**Figure 38:** MR leave-one-out plot IQ on Schizophrenia

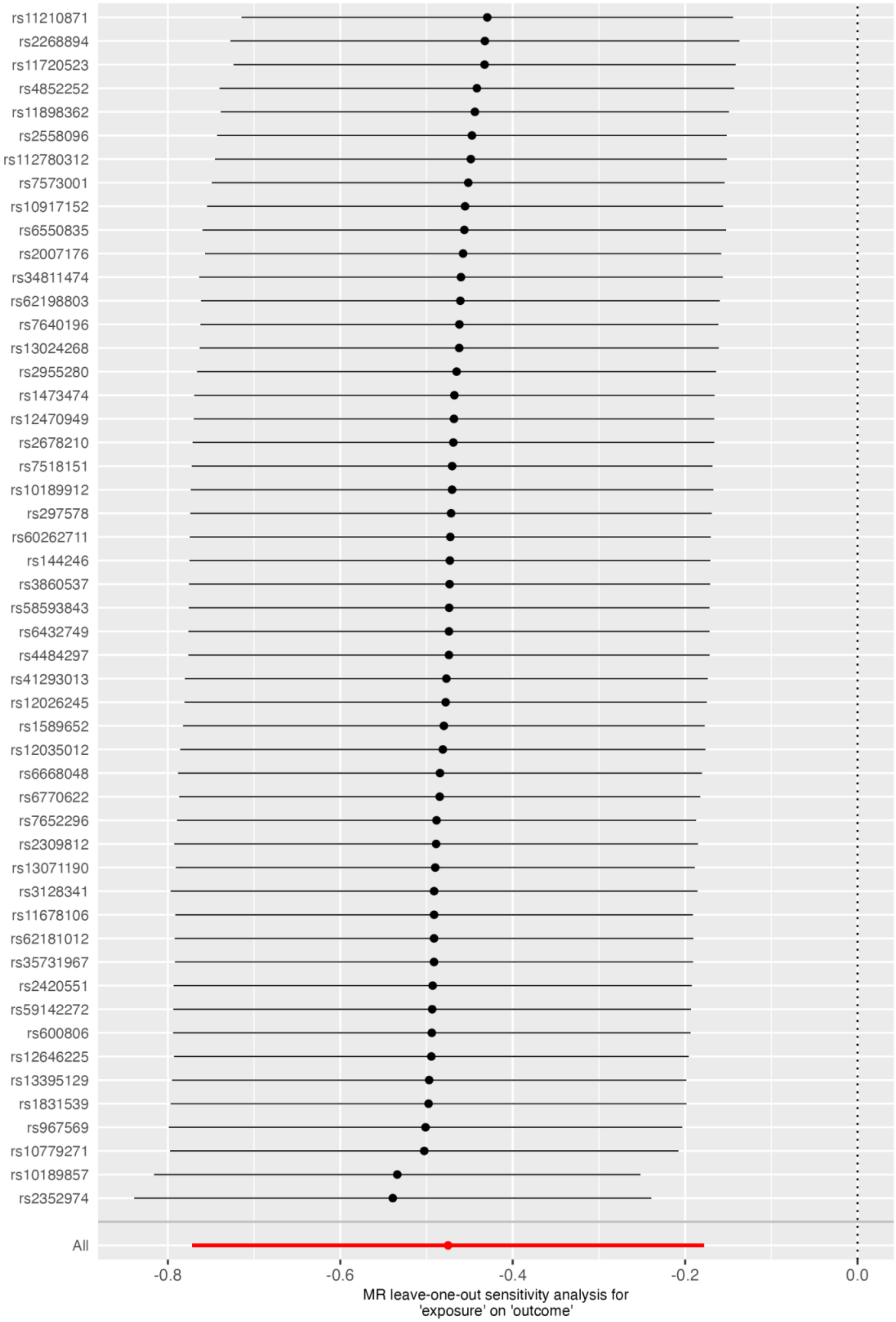

**Figure 39:** MR scatter plot IQ on Bipolar

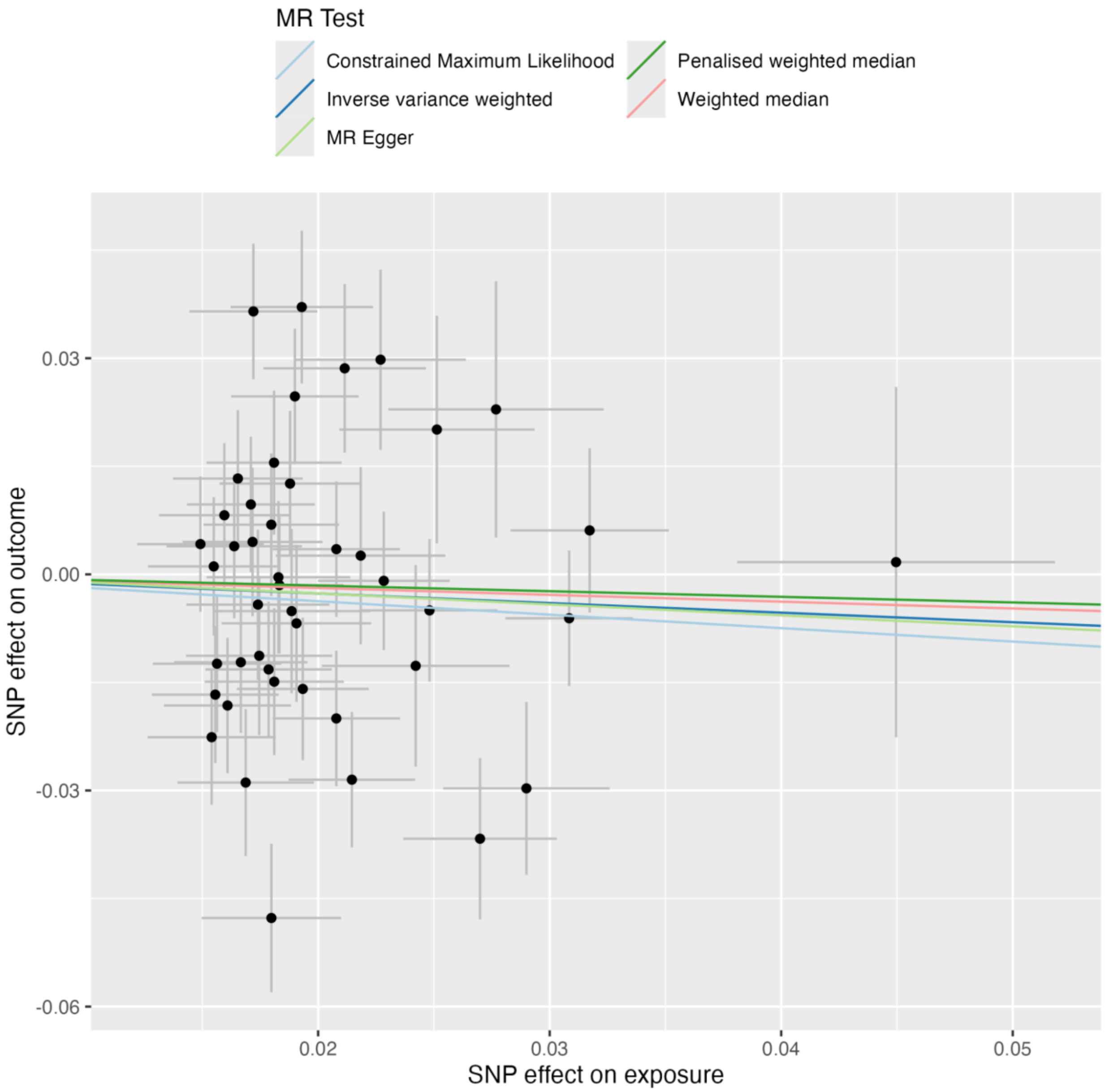

**Figure 40:** MR leave-one-out plot IQ on Bipolar

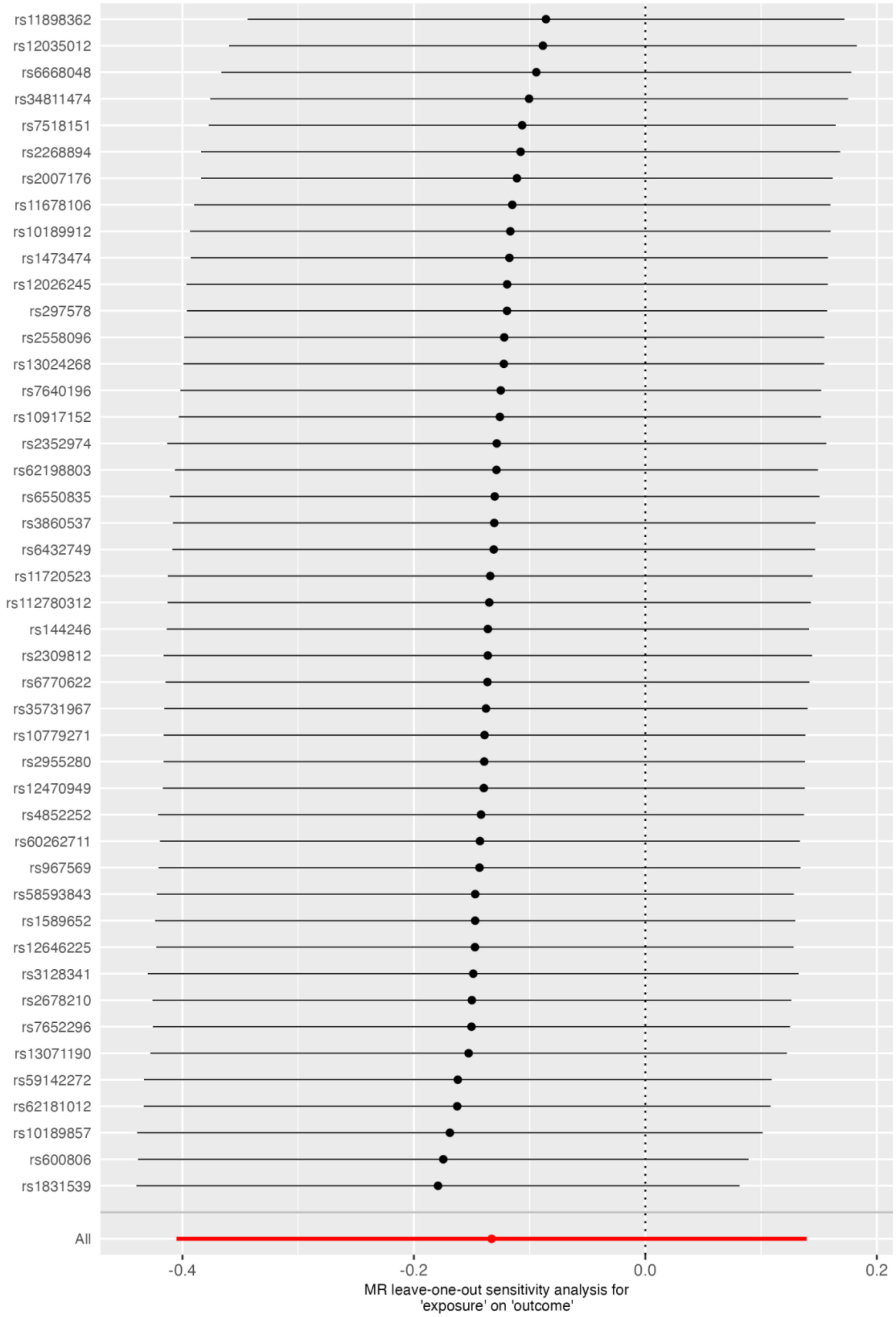

**Figure 41:** MR scatter plot IQ on SZspecific

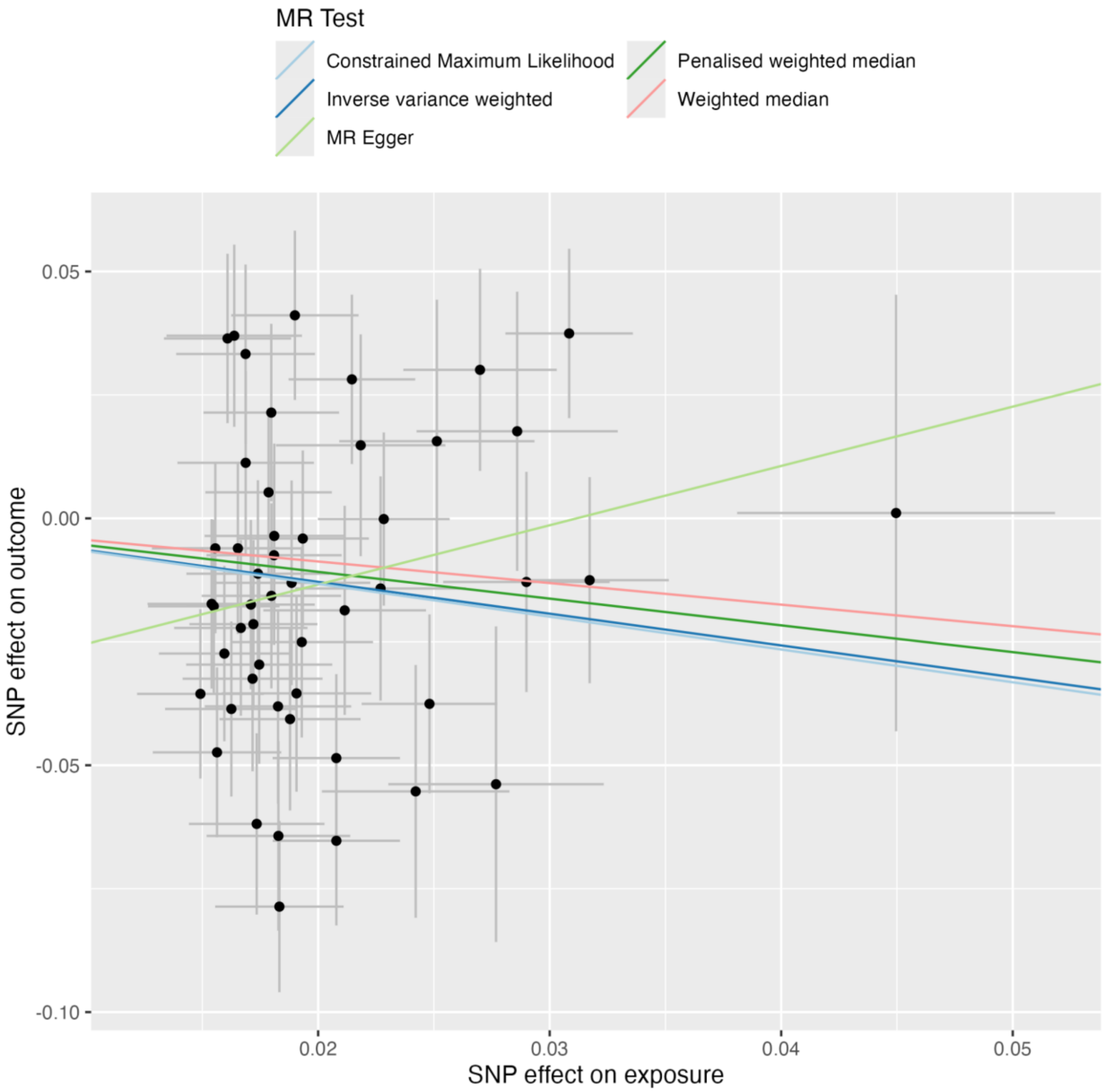

**Figure 42:** MR leave-one-out plot IQ on SZspecific

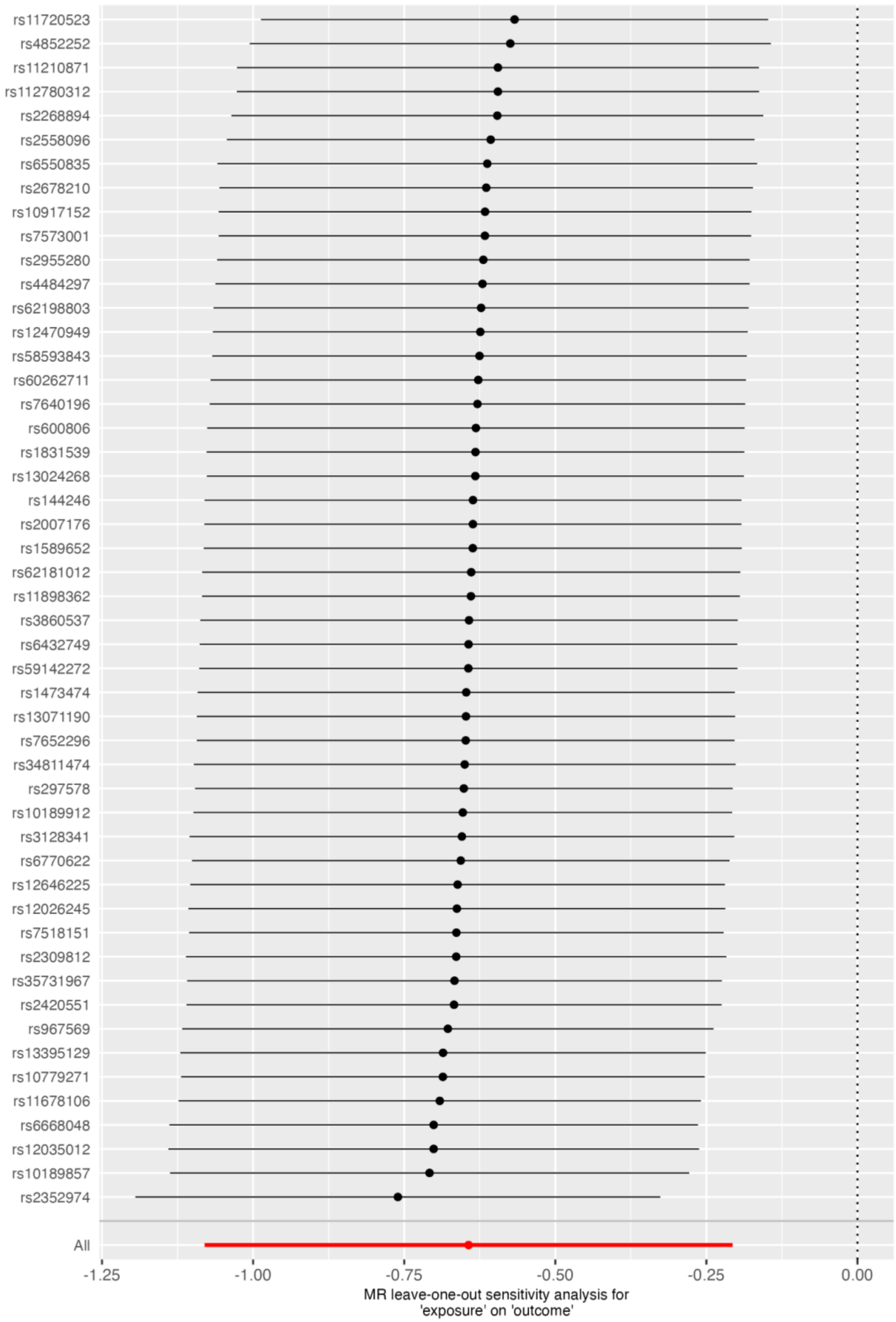

**Figure 43:** MR scatter plot IQ on PSYshared

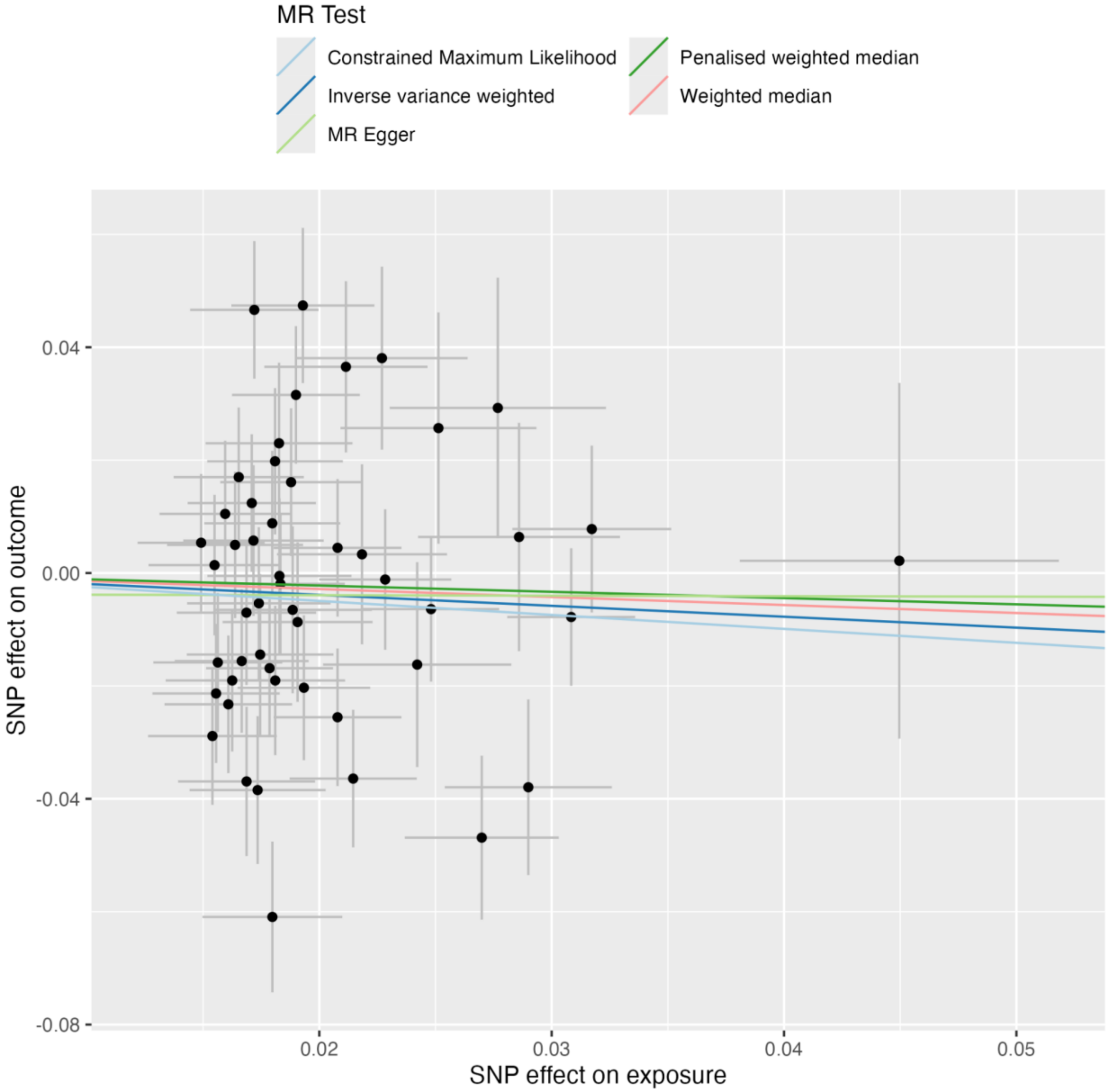

**Figure 44:** MR leave-one-out plot IQ on PSYshared

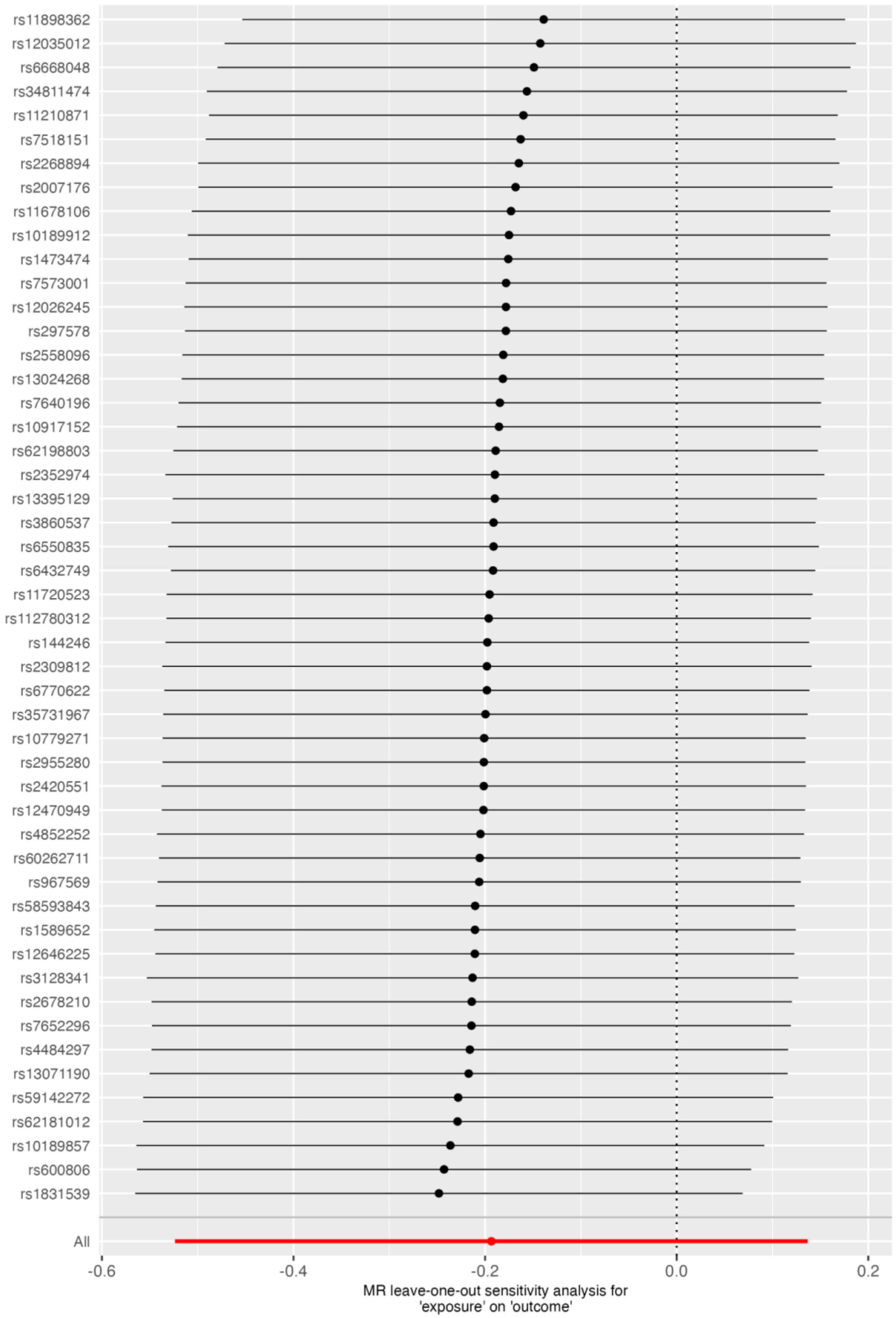
